# Village-scale persistence and elimination of *gambiense* human African trypanosomiasis

**DOI:** 10.1101/19006502

**Authors:** Christopher N. Davis, Kat S. Rock, Erick Mwamba Miaka, Matt J. Keeling

## Abstract

*Gambiense* human African trypanosomiasis (gHAT) is one of several neglected tropical diseases that is targeted for elimination by the World Health Organization. Recent years have seen a substantial decline in the number of globally reported cases, largely driven by an intensive process of screening and treatment. However, this infection is highly focal, continuing to persist at low prevalence even in small populations. Regional elimination, and ultimately global eradication, rests on understanding the dynamics and persistence of this infection at the local population scale. Here we develop a stochastic model of gHAT dynamics, which is underpinned by screening and reporting data from one of the highest gHAT incidence regions, Kwilu Province, in the Democratic Republic of Congo. We use this model to explore the persistence of gHAT in villages of different population sizes and subject to different patterns of screening. Our models demonstrate that infection is expected to persist for long periods even in relatively small isolated populations. We further use the model to assess the risk of recrudescence following local elimination and consider how failing to detect cases during active screening events informs the probability of elimination. These quantitative results provide insights for public health policy in the region, particularly highlighting the difficulties in achieving and measuring the 2030 elimination goal.

**Author summary:** *Gambiense* human African trypanosomiasis (gHAT) is a vector-borne infectious disease that causes sleeping sickness across many African countries. Reported gHAT cases show a continued decline, but it is unclear if this is sufficient to reach the WHO goal of stopping transmission by 2030. We develop a stochastic model necessary to address the critical question of persistence of gHAT infection at the local-scale. In contrast to other commonly studied infections, we predict long-term persistence of gHAT in small populations (< 1, 000 people) despite very low prevalence. Our local-scale predictions (together with previous larger-scale studies) suggest that, to achieve regional elimination, controls need to be widespread and intensified in the worst affected regions, while the movement of infected people could rapidly lead to re-emergence.

## Introduction

*Gambiense* sleeping sickness (*gambiense* human African trypanosomiasis, referred to here as gHAT) is a tsetse-borne neglected tropical disease (NTD) caused by the parasitic protozoa, *Trypanosoma brucei gambiense*. There has recently been a decline in global cases, with just 1,420 cases reported in 2017, compared to 10,466 reported in 2007 [1]. This decline is largely attributed to improvements in the active screening and treatment campaigns that have been carried out in many regions [2]. In 2012, the World Health Organization (WHO) set targets for elimination of gHAT as a public health problem; these were updated in 2017 to: reducing the area at risk of reporting more than 1 case per 10,000 people per year by 90% as compared to the baseline for 2000–2004 [2], and for fewer than 2,000 reported cases per year, by 2020 [3]. There is also a more stringent goal of stopping transmission of gHAT by 2030 [3].

Cases of gHAT primarily occur in West and Central Africa, but the distribution of infection is heterogeneous, with highly clustered incidence resulting in disease foci [4, 5]; reported prevalence often varies greatly over short distances, even between neighbouring villages [6]. This local variation suggests there is a complex spatial structure to the infection. With the observed global decline in reported cases and with many (but not all) foci likely to achieve less than 1 case per 10,000 people by 2020 [7, 8], it is increasingly important to understand both where the disease is most likely to persist and why this might be the case.

In regions where gHAT cases are no longer observed (and where local elimination has been achieved), it is possible for the disease to be re-introduced through movement of either infected humans or infected tsetse, and it may become re-established especially if active case-finding has not been maintained [9]. It is therefore clear that, while the current active screening is highly successful in many regions, an understanding of stochastic re-invasion and re-establishment in local populations is also essential to guide post-elimination policy planning.

There are two stages of gHAT, with Stage 1 following initial infection and Stage 2 defined after trypanosomes have crossed the blood–brain barrier [10]; these stages currently require very different treatments. Patients are hospitalised for the treatment duration and are advised to recover at home afterwards, on average, for a total time of 6 months [11]. Without the treatment, individuals typically progress from Stage 1 to Stage 2 after 18 months and most would likely be expected to die from meningoencephalitis after approximately 3 years [12].

Active detection of gHAT occurs through population-level screening, which is implemented in many endemic regions by mobile teams travelling to settlements and testing the available population [6]. Teams are generally able to screen substantial proportions of the local population (often over 70%) [13]; however, some socio-demographic groups (notably working adult males) frequently do not present for testing. Previous work has indicated that the individuals missed in screening are also more likely to be more highly exposed to tsetse bites [14, 15], potentially due to working in tsetse-infested forested and riverine areas. Such high risk core-group individuals pose a barrier to elimination [8, 16–18]. In parallel with active screening, passive detection occurs when individuals voluntarily attend medical facilities for testing, usually after the onset of more significant symptoms, and are thus most often in Stage 2 of the disease [11].

Mathematical models of gHAT have been beneficial in identifying the effectiveness of differing control strategies and predicting when elimination is likely to occur [19].

However, much of the modelling work on the gHAT infection dynamics has been done in large populations using deterministic models, either for an entire regional infection focus or at a health zone level (approximately 100,000 people) [8, 15, 17, 20, 21]. Here we translate the deterministic model of Rock et al. [17] to a stochastic framework, designed to capture the infection dynamics and chance extinction at the village-scale. As such, our model is mechanistic and so captures details of the biology and epidemiology, allowing modification of model components to predict a number of different scenarios and control options.

Much of the previous work on the stochastic persistence of infection has tended to focus on measles in developed countries [22, 23]. Measles is directly transmissible, has a high reproductive ratio (12–18 compared to approximately 1–1.1 for gHAT [1]) and a high incidence before immunisation programs were introduced; yet, in contrast to gHAT, measles only persists in large populations of above approximately 300,000 and even then relies on frequent reintroductions [22, 23].

We use our model to address the dynamics of gHAT in the villages within the Yasa-Bonga and Mosango health zones of the DRC (Fig 1); a region that has an extremely high incidence of gHAT. The model has already been fitted to the available epidemiological data from the WHO HAT Atlas (Fig 1) [7, 13], with parameters inferred from regional reporting patterns and screening mimicking observed village-scale records (see Methods and S1 Appendix).

**Fig 1.**
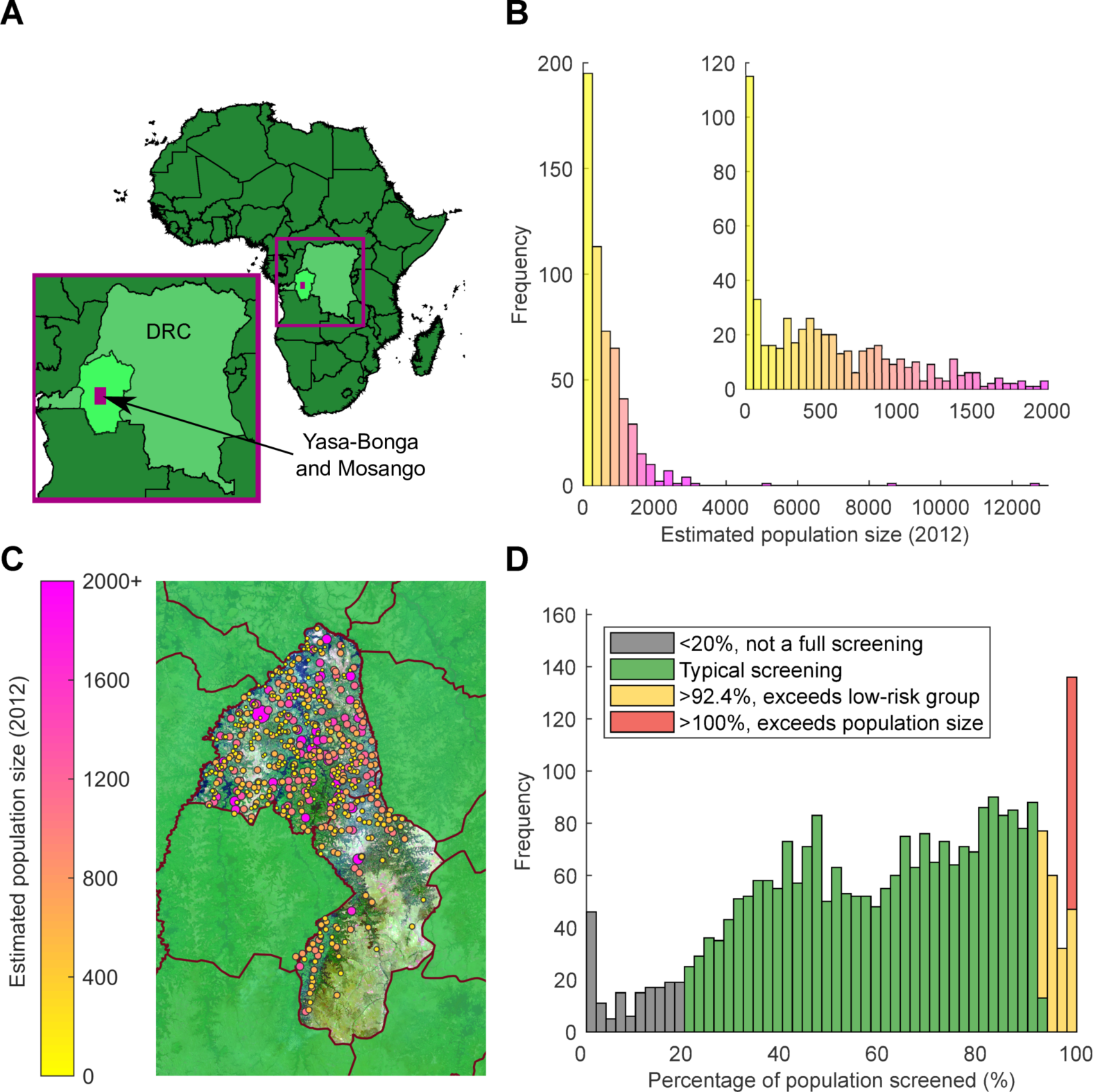
Visualisation of WHO HAT Atlas data for the study health zones: Yasa-Bonga and Mosango. (A) Map of Africa, showing the DRC (lighter green) with former Bandundu province highlighted in the lightest green. The area containing the health zones Yasa-Bonga and Mosango (presented in more detail in Fig 1C) is covered in a purple box. (B) Histograms of the estimated population sizes of villages in the region for 2012. The inset figure highlights the distribution of population sizes less than 2,000 individuals. Population sizes have a range of 3 to 12,645. (C) Detailed map of locations of settlements within the study region (Fig 1A, purple box), colour and radius of the circle represent population size of the individual settlements. The satellite image shown for the Yasa-Bonga and Mosango health zones is from Landsat-8 accessed through https://earthexplorer.usgs.gov/ from the U.S. Geological Survey. (D) Histogram of the coverage achieved in visits to settlements as part of the active screening programme. When annual screenings are larger than the estimated accessible population (yellow and red bars), this may indicate either multiple screenings in a given year, or misreporting, when individuals are attending from neighbouring settlements.

## Methods

### Data

The Democratic Republic of Congo (DRC) has the highest burden of gHAT cases (1,110 cases out of 1,420 reported globally in 2017 [1]), and 46% of these cases are concentrated in the former Bandundu province [24]. We focus on models and data for two high-prevalence health zones in this former province (now in Kwilu province): Yasa-Bonga and Mosango. Reported case information made available by the WHO HAT Atlas [7, 13] details the locations of settlements with estimates of population size, the years that active screenings took place, the number of people screened and the resulting newly identified gHAT cases. In Yasa-Bonga and Mosango, we consider 559 settlements, which experienced 2,701 active screenings in 2000–2012, each where a full village population was targeted to be screened, resulting in 4,875 detected gHAT cases, as well as 2,496 additional cases from detected by passive surveillance. Our model utilises these recorded screening patterns to simulate interventions, while the active and passive cases detected in each settlement are stochastically generated by the model.

Population sizes for settlements are also obtained from census estimates within the WHO HAT Atlas (Fig 1B) [7, 13]; we account for an estimated yearly population growth of 2.6% [25]. Screening coverage from active surveillance is then calculated as the number of individuals screened divided by this population estimate in each year.

Annual screenings larger than the estimated accessible population (yellow and red bars in Fig 1D) may indicate either multiple screenings in a year or misreporting of individuals attending from neighbouring settlements. Low annual screening coverage (at less than 20% of the population size, gray bars in Fig 1D) is assumed to represent individuals screened outside their home settlement and therefore is not considered as a complete active screening of any given village.

All relevant model data are displayed within the paper and the Supporting Information files (S1 Appendix, Table S2). Epidemiological data for the study were provided by the WHO in the frame of the Atlas of gHAT which may be viewed at www.who.int/trypanosomiasis_african/country/risk_AFRO/en and may be requested through Jose Ramon Franco.

### Modelling

The infection dynamics are described by a stochastic compartmental Ross–Macdonald-type model [26–29] extended from the previous work of Rock et al. [17] (using the formulation of Model 4 from that study) (S1 Appendix, Fig S1). The model captures a population of humans, which is initially partitioned into those at high and low risk of being exposed to tsetse bites. Each person will transition between five different epidemiological compartments: susceptible; exposed (or latent); Stage 1 infection; Stage 2 infection; and hospitalised (and temporarily removed). We assume that there is natural mortality from all compartments, which leads to replacement of that individual as a susceptible in the population.

The risk structure is used to capture the behaviour of the small proportion of individuals that both work in the habitat of many tsetse and so have a higher biting exposure and also do not partake in active screening, thereby acting as a human reservoir of infection [16–18]. The proportion of the individuals in the high risk group is estimated through extensive model fitting to be 7.6% of the population for these health zones [17]. While the high risk group have a higher biting exposure, the probability of tsetse infection per single infective bite is the same for both risk groups. This model structure, where individuals are either high risk and non-participating in active screening or low risk and randomly participating in active screening, was selected using the deviance information criterion (DIC), which assigns low scores for models with high posterior mean log-likelihood and penalises models with more parameters.

Tsetse in the model are similarly compartmentalised into four epidemiological states: teneral (unfed); non-teneral yet uninfected; exposed (or latent); and infected. The distinction between teneral and non-teneral yet uninfected is used to capture the observation that tsetse are far more susceptible to infection at their first blood meal than at any subsequent blood meals [30]. The effect of a possible animal reservoir is not considered, since its role remains unclear [15, 17, 20, 31, 42] and its inclusion does not significantly improve the match between model outputs and currently available data in this setting [17].

Additional to the epidemiological and demographic processes, we simulate the effect of active screening and passive detection of cases. Passive detection (and disease-induced mortality) is assumed to occur at a fixed (per capita) rate for all Stage 2 infected individuals [11]. Active screening takes place annually; the proportion screened is either replayed from the historic pattern for that settlement or chosen randomly from the set of all screening coverages recorded, allowing a greater range of scenarios to be explored. Since Yasa-Bonga and Mosango are high endemicity regions, we assume that the screening coverage and frequency remains constant over time but note that these quantities are somewhat affected by population size (S1 Appendix, Fig S8).

Individuals in the low risk group are selected randomly for screening, irrespective of epidemiological status, and those that are found to be infected are moved to the hospitalised class. We assume that screening only applies to low risk individuals, such that screening coverages greater than 92.4% (the estimated proportion in the low risk group) [17] are truncated (Fig 1D). In the field, the diagnostic process is complex and multi-stage [32]; however, in the model, we collapse this into characteristics for the whole algorithm, which is assumed to be 91% sensitive [33] but 100% specific. False negatives remain undetected in the settlement, but by assuming 100% specificity, there are no false positives as we assume confirmation by microscopy will be carried out due to the low case numbers.

For the majority of this paper, we model the dynamics as a closed population, without emigration or immigration, so that once the disease has gone extinct in a population it cannot be re-introduced. This removes a critical dependency in model formulation, and greatly simplifies the presentation of results. In reality, no population is ever completely isolated; however we show that the expected rate of infectious imports is very low and does not affect our main results or conclusions (S1 Appendix).

### Parameters

Model parameters for the underlying compartmental model are taken from the previous work of Rock et al. [17]. The values of these parameters were taken from literature, where well-defined, and otherwise inferred by model fitting using a Metropolis–Hastings MCMC algorithm, which sought to match the deterministic model to observed cases and screening effort in Yasa-Bonga and Mosango. The values used in this manuscript are the median of the distributions inferred using MCMC methodology applied to the aggregate annual data from Yasa-Bonga and Mosango. Therefore, the parameter values are specific to the study region, and the model is well-matched with the incidence data from active and passive surveillance. A full list of parameters is given in the S1 Appendix, Table S1.

## Results

We use our model to address the dynamics of gHAT in the villages within the Yasa-Bonga and Mosango health zones of the DRC (Fig 1), a region that has an extremely high incidence of gHAT. Using the estimated population sizes and the reported levels of active screenings (Fig 1B and D), we validate the stochastic model by comparing the observed and predicted screenings that did not detect any cases (which subsumes both local extinction and failure to detect). We then consider the probability of local gHAT persistence across a range of population sizes and different control scenarios, as well as the probability of re-invasion. Finally, we focus on whether not detecting cases in a series of active screens can inform on whether local elimination has been achieved, noting that WHO guidelines suggest annual active screenings until there have been three consecutive years of no new cases, followed by a further screening with no cases after three years [32].

### Comparison with data

While the underlying deterministic model has been fitted to the aggregate data from this region, it is important to assess the behaviour of the stochastic model against village-scale observations. Unfortunately, local disease extinctions cannot be directly observed; failure to discover any cases does not necessarily mean that the infection is not present, simply that it has not been detected. Thus, to validate our model, we make comparisons between the simulated predictions and WHO HAT Atlas data [7, 13] for the probability of detecting no cases on an active screening (termed zero-detections for brevity), which is a combination of failure to detect and local extinction. We compare model predictions to observations by calculating the percentage of zero-detections in aggregations of 100 active screenings with similar village population sizes (Fig 2A). We find very strong agreement between model predictions and data, with a pronounced decline in zero-detections for larger populations.

**Fig 2.**
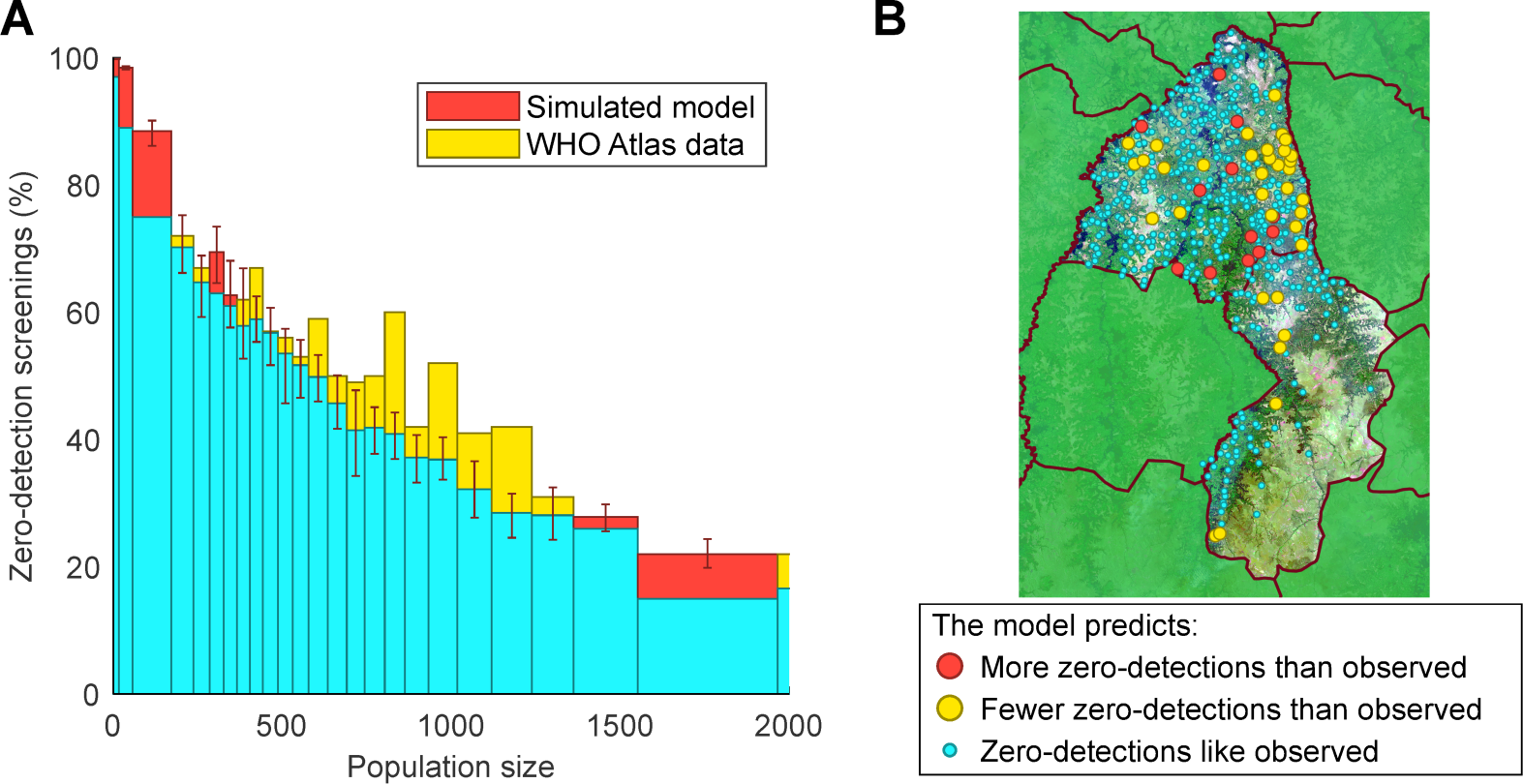
Comparison of model predictions and data for active screenings with no detected cases (zero-detections). (A) Histogram by population size of the percentage of active screenings that find no new gHAT cases for both the model and the HAT Atlas data. Each bar represents 100 screenings of simulated results (averaged over 10,000 replicates) from the model that uses the observed pattern of screenings and compares to the data. Values where the model predictions have more zero-detections than the data are in red, while the reverse is shown in yellow. Error bars represent the 95th percentile of model results. (B) Map of populations in Yasa-Bonga and Mosango showing the settlements with significant differences (at the 95% level) in the expected proportion of active screenings with no cases detected. Red circles are where the observed number of active screenings with zero-detections is below the 95th percentile of the model; yellow circles are where the data falls above the 95th percentile; small blue circles are for data that lie within the 95th percentile of predictions and therefore are well described by the model. The satellite image shown is from Landsat-8 accessed through https://earthexplorer.usgs.gov/ from the U.S. Geological Survey.

For individual settlements, those where the number of zero-detections lie outside the 95th percentile of model predictions are notably spatially clustered (Fig 2B). In 2.1% of settlements (red), there are significantly fewer zero-detections than predicted and hence greater persistence; these villages are generally localised around the main river through the region. In 6.3% of settlements there are significantly more zero-detections than expected (yellow), and these are clustered far from the major rivers and in upland areas. Since tsetse are most densely distributed surrounding riverine areas [34], this spatial clustering may indicate the need for spatially heterogeneous parameters that reflect the suitability of the local environment for tsetse. However, given that more than 90% of villages fall within our prediction intervals, we believe the homogeneous parameters capture the general stochastic behaviour of this region.

### Local gHAT persistence

Local gHAT persistence, where human infection is maintained in a settlement, is affected by many factors (Fig 3), including the population size of the settlement; the vector-to-host (tsetse-to-human) ratio; the exposure to the tsetse; the screening procedure; and any movement of infected individuals between populations. We calculate the probability of persistence by stochastically simulating the epidemic for 16 years from the endemic (uncontrolled) disease equilibrium. If there is zero gHAT infection in the human population after a given number of years and no further human infection emerges in the following year from infected tsetse, we say in this simulation there is local disease elimination with no immediate threat of re-emergence (Fig 4A provides a justification for this criteria). This procedure is repeated for multiple population sizes; the proportion of simulations that retain infection after a given number of years determines the probability of persistence (Fig 3).

**Fig 3.**
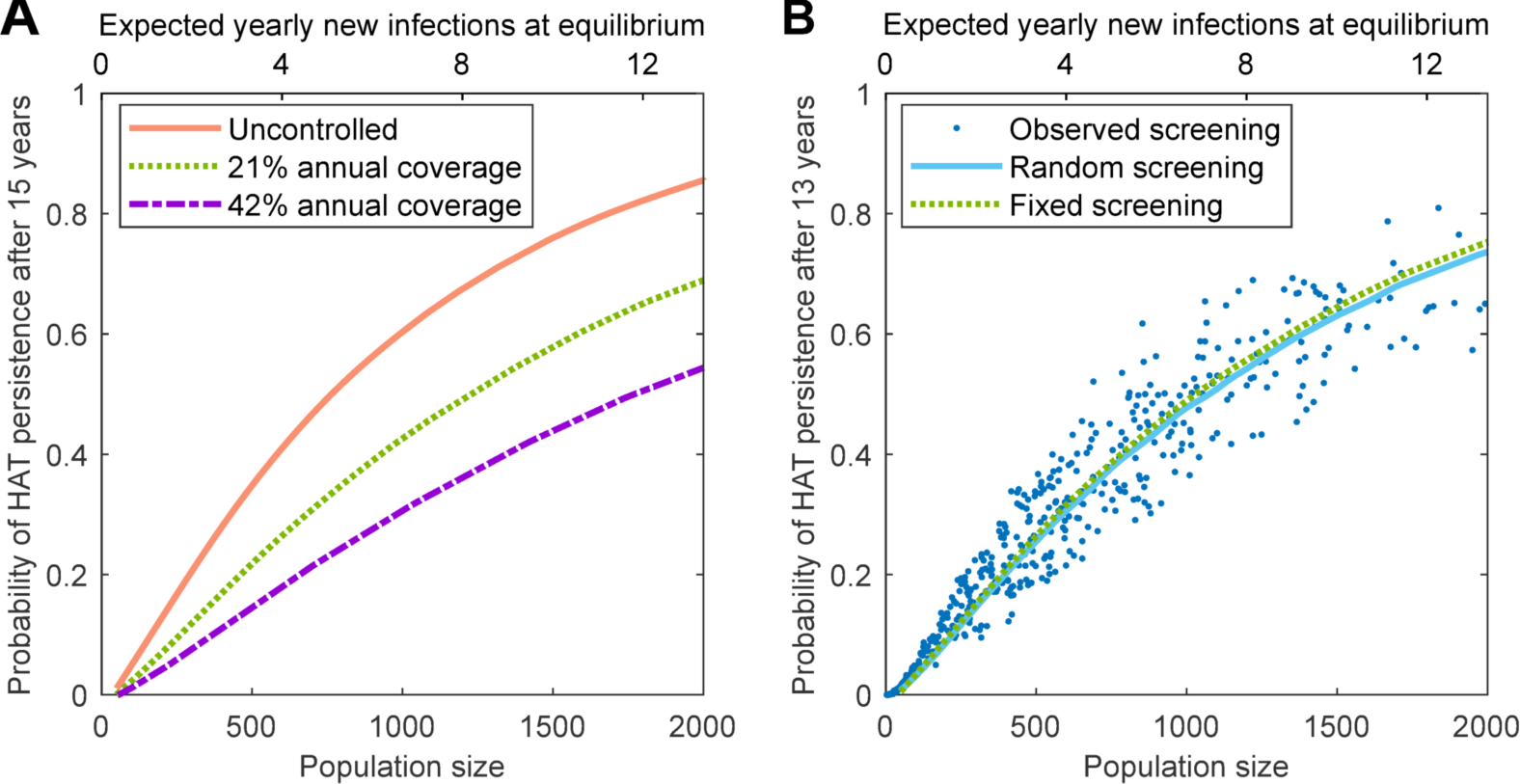
Predicted probability of gHAT persistence in isolated settlements. Simulations are started at the endemic (uncontrolled) equilibrium and iterated forwards (without infectious imports), while the persistence of infection is recorded. This is repeated 100,000 times for settlement population sizes between 50 and 2,000 individuals. The expected number of yearly new infections if the system were at equilibrium is proportional to the population size and is given by the top scale. (A) Impact of active screening on gHAT persistence; annual screening at a fixed coverage per year yields a drop in persistence with increased coverage. (B) Comparison of screening assumptions on the persistence of gHAT. The solid curve shows results where annual screening coverages were randomly sampled from all observed coverages; dots represent the individual settlements recorded in the WHO HAT Atlas for Yasa-Bonga and Mosango health zones [7, 13], where the reported coverage in each year is used. There were sufficient simulations such that confidence intervals are too small to be visible.

**Fig 4.**
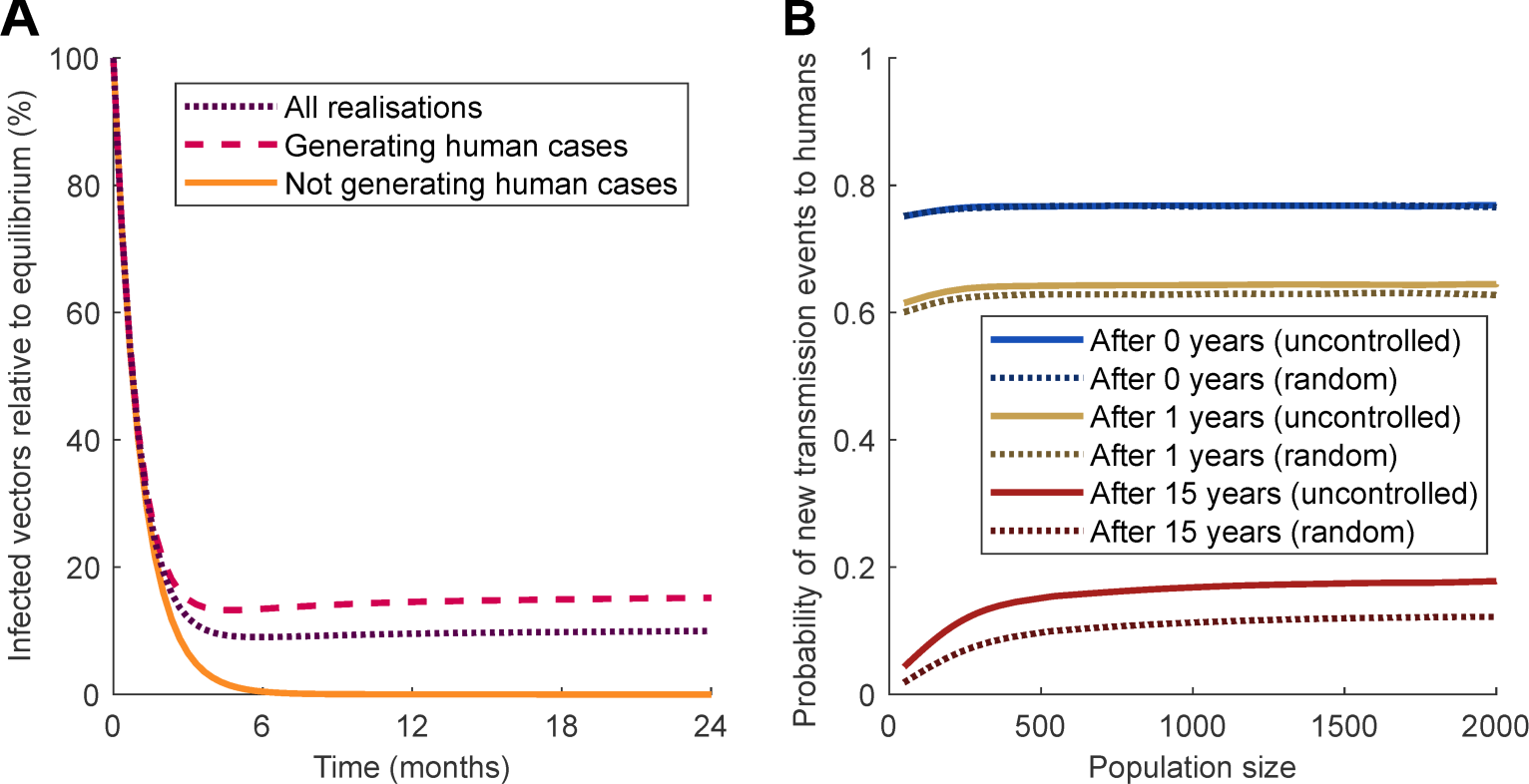
Dynamics of extinction and reintroduction. (A) Starting with no infected humans but the tsetse population at its endemic equilibrium and a settlement size of 1,000 individuals, the model predicts a dramatic decline in the infected tsetse population, depending on whether subsequent human cases are generated by the infected tsetse. (B) Extending this model further for a range of initial conditions, we examine the probability that at least one human case is generated and the infection persists for a given time, when starting with one infected human and no infected tsetse for both uncontrolled and random screening.

We focus on settlements with fewer than 2,000 inhabitants, typical of Yasa-Bonga and Mosango (Fig 1B), and use regionally specific parameters. In all scenarios investigated, we find that persistence increases with increasing population size (Fig 3). This echoes results from other infections [22, 23, 35, 36], where small populations with low incidence experience a greater impact of stochasticity and chains of transmission that are more likely to be broken. In addition, given that long-term persistence relies on persistence for shorter time intervals, the probability of persistence decreases with time (S1 Appendix, Fig S9B–C). The relatively long persistence times of gHAT, compared to the frequently studied persistence of childhood diseases [22, 23], are attributable to the long time-scale of gHAT infection in the absence of active interventions [37].

The addition of active screening (leading to the treatment of detected cases) decreases the probability of persistence across all population sizes, since removing infected individuals leads to a greater chance of breaking chains of transmission (Fig 3A). Increasing levels of screening, beyond the observed 21% average, leads to further reductions for persistence. We compare three assumptions for active screening (Fig 3B): that each population is screened annually at a fixed coverage equal to the regional average (21%); that each population experiences screening coverages sampled from the regional pattern including not screening in a given year; and simply replaying the recorded pattern of active screening in each village. Despite the very different distributions of screening effort, all three of these assumptions produce comparable levels of persistence.

### Re-invasion probability

Following localised elimination of infection, populations remain vulnerable to re-invasion; we investigate the potential for re-establishment of sustained transmission in a settlement for different invasion scenarios. For a village of 1,000 individuals, following the elimination of infection in humans, the level of infection in tsetse falls rapidly, even when starting at the endemic level in these vectors (Fig 4A). In approximately 65% of simulations, the initially infected tsetse generate human cases, and the level of infection in vectors rapidly plateaus; otherwise, infection is eliminated from the location within six months due to short vector life expectancies in comparison to the human hosts. This validates the previous simulation assumption (Fig 3) that local disease elimination can be considered achieved after a period of one year in which there are no infected humans, as the number of infected tsetse will also become negligible.

The probability that a population of tsetse, infected at the endemic equilibrium (0.02% of tsetse exposed or infected), will lead to re-establishment of infection is predicted to be a function of settlement size (S1 Appendix, Fig S10A). Small populations are unlikely to see any new human infections, and those that are generated fail to persist. However, even for large population sizes of 2,000 individuals, the chance of continued transmission beyond one year is only 55% and is less than 10% over 15 years. For lower levels of infection in the tsetse population, the risk of successful re-establishment is proportionally reduced.

In contrast, if re-invasion of an infection-free population is due to the movement of an infected person into the settlement (in the absence of infected tsetse) the probability of re-establishment over different time-frames is largely unaffected by the population size (Fig 4B). We predict a high probability (> 70%) of short term re-invasion, but a more limited chance (< 20%) that this will generate persistent infection for 15 years or more. This is typical of stochastic dynamics of infection with low basic reproductive ratio (*R*_0_), where, although short chains of transmission are likely, it is difficult for the infection to fully establish.

### Detecting elimination

As discussed above, zero-detections may be an indication that there is no infection in a settlement; however, it may simply be that infected individuals were not screened or were false negatives. Despite this, there is a temptation to associate zero-detection with zero infection; we therefore use our simulations to tease apart this complex relationship. The model replays the observed pattern of screening in each settlement, but we perform multiple simulations to ascertain the probability that infection has been locally eliminated following one, two or three consecutive zero-detection screenings, in which at least 20% of the population are screened (Fig 5). We also insist that no passive cases were detected between the screening events. Our standard assumption, in agreement with parameter inference for this region, is that 26% of Stage 2 infections, where people either self-report or likely die of gHAT infection, are detected and reported [17], but we also investigate 0% (no passive reporting) and 100% (all passive cases and deaths are reported) for comparison.

**Fig 5.**
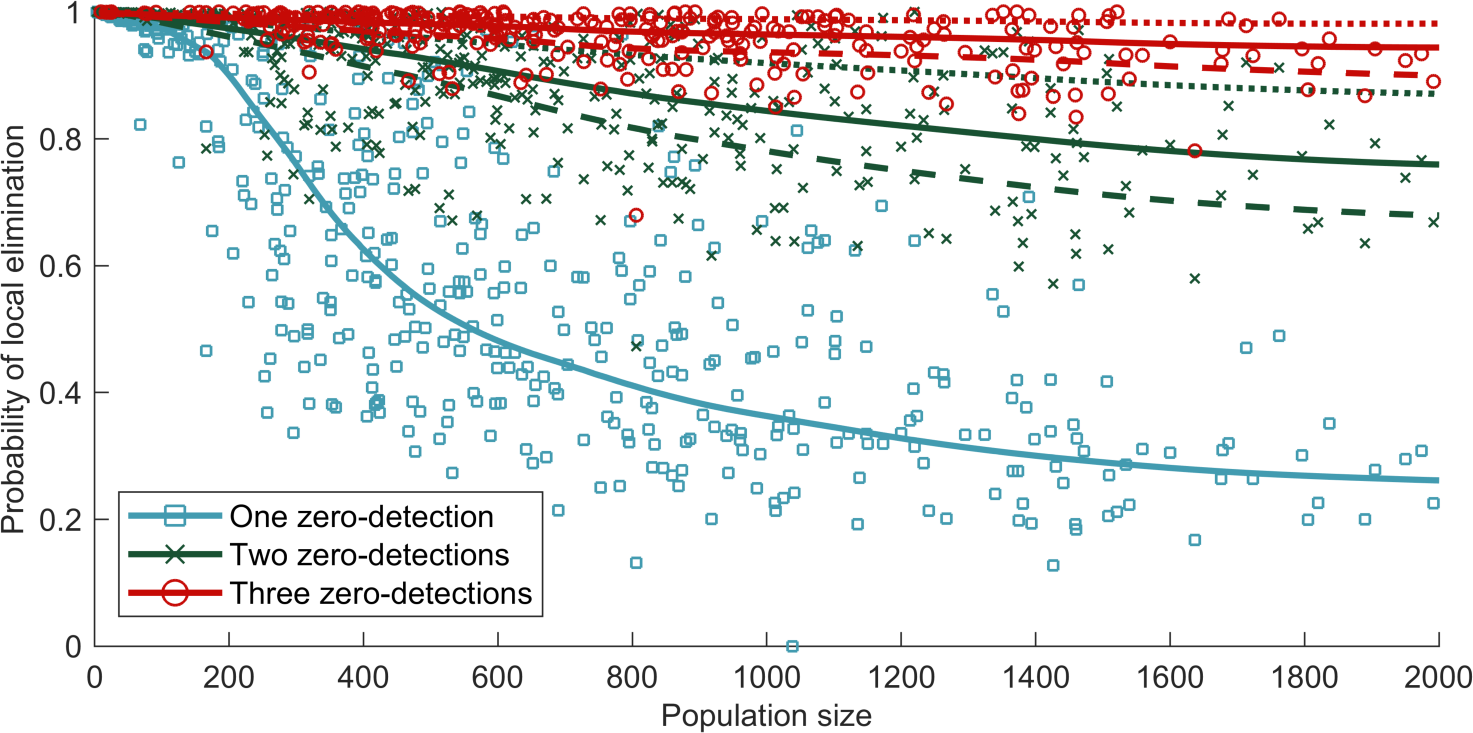
Probability of elimination in a settlement, given consecutive zero-detections with no detected passive cases. (A) Consecutive zero-detections means consecutive in the observed years of screenings, not necessarily in consecutive years, and with no passive cases detected in between. Each point represents the average from multiple simulations of individual settlements where the reported pattern of screenings is replayed. The points and solid lines assume a reporting rate of 26% [17], while the dashed and dotted lines show reporting rates of 0% and 100%, respectively. Sufficient simulations are used such that the confidence intervals are small (unobservable on the scale of this graph). Lines represent a weighted local regression fit. Active screenings where fewer than 20% of the population are assessed are excluded from our analysis due to the small sample sizes (alternative cut-offs of less than 10% and less than 50% are presented in the S1 Appendix, Fig S12).

For small settlements, given that long-term persistence is unlikely (Fig 3), even a single zero-detection screen (Fig 5, blue) is frequently associated with local elimination. For larger populations, a single zero-detection has limited predictive power, and three consecutive zero-detections are needed to have any degree of confidence, in which case the reporting of passive cases plays a noticeable role. There is, however, significant variation between settlements, reflecting very different patterns of reported screening. Moderate population sizes of between 200 and 1,000 individuals show extreme variation in the ability to predict local elimination, while smaller villages have less variation, in part due to rarely being screened before 2009 (S1 Appendix, Fig S8C).

### Importation rates

Regional persistence relies on more than independent persistence in individual settlements. It is likely that the occasional movements of infected people lead to a stochastic meta-population paradigm [41], where rare local extinctions of infection are balanced by external imports. However, the agreement between model and data (Fig 2), together with the low prevalence of infection, indicates that imports are likely to be rare.

We make this more quantitative by fitting an importation rate of infection, proportional to the size of the population. Before interventions, the presence/absence of infection in a population reflects the equilibrium balance between extinction and re-colonisation; we can therefore use the presence of infection at the first recorded active screening within a village as measure of the equilibrium state. Fitting the external importation rate to the probability of detecting infection in a village at the first active screen gives the best fit when the rate is small at just 3.4×10^−6^ per person per day (Fig 6A). Moreover, it is assumed that the importation rate declines over time as the overall prevalence in DRC reduces.

**Fig 6.**
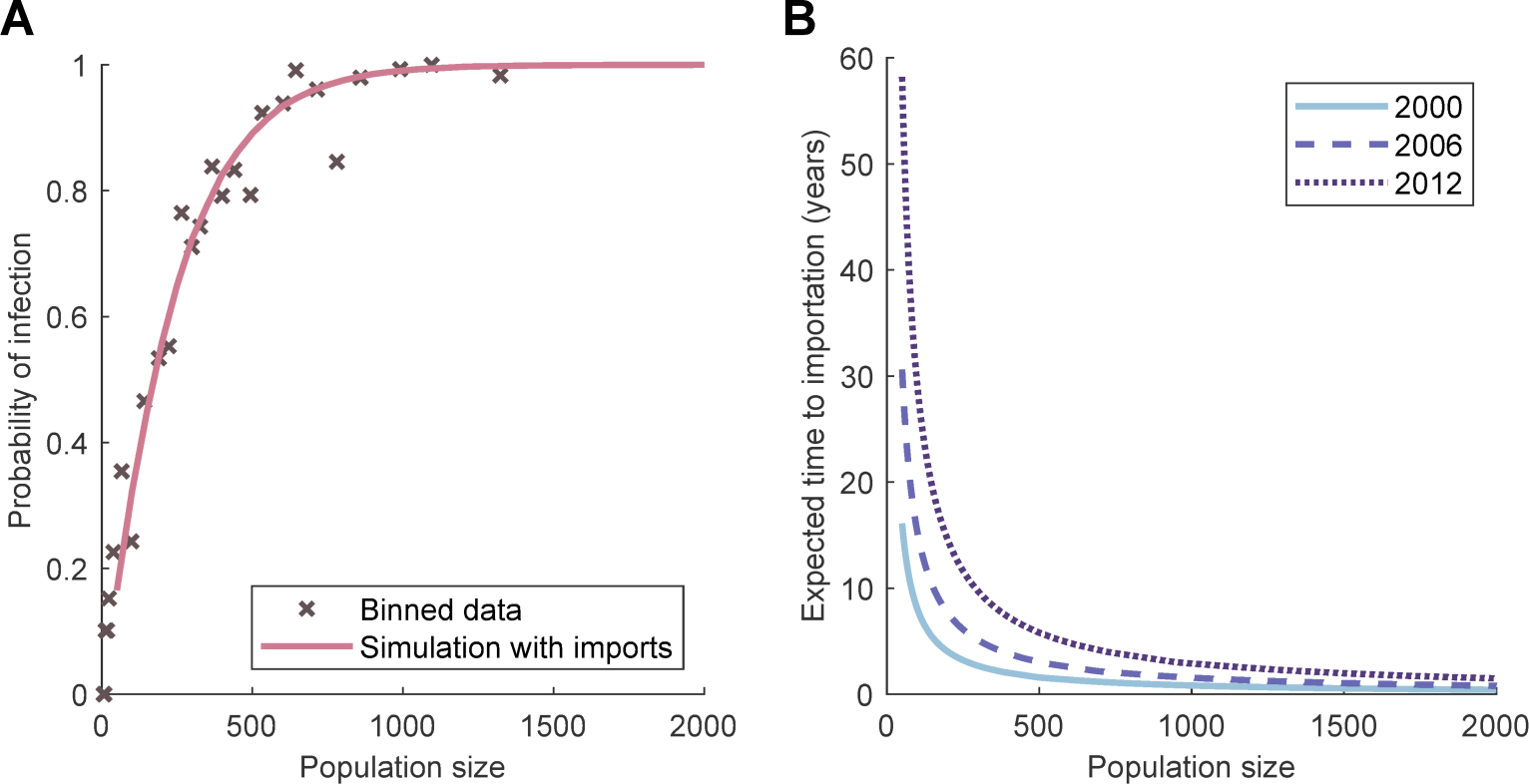
Simulating external importations of infection into village populations. (A) By running simulations with different values for the external infection parameter, we find the best fit — to data binned by population size from the WHO HAT Atlas on whether there are any detected cases on the first active screening — is when imports per susceptible individual are equal to 3.4 ×10^−6^ days^−1^ (solid line). (B) Curves of the expected time for an external importation into a village population using the fitted importation parameter. The importation parameter is assumed to decay at the same rate as total number of cases in time in the DRC (see S1 Appendix, Fig S8A).

For most population sizes in the region, the expected time to importation is therefore relatively long (Fig 6B), and in many cases a single importation will not cause further transmission events (Fig 4), leading us to conclude that in general the level of importation will not qualitatively change our results. This is made more explicit in the supporting information (S1 Appendix, Fig S6), where it is shown that the model with the fitted level of infectious imports generates comparable results to those described in the main paper. This justifies our modelling assumptions that villages act as isolated populations and that importation of infection is unlikely to perturb the dynamics; instead, we are able to separate the processes of local elimination and re-invasion.

## Discussion

Despite global declines in reported cases over the last decades, *gambiense* gHAT remains a problem in many focal areas [38]. These regions, concentrated primarily in the DRC, represent a significant challenge to achieving the WHO 2020 and 2030 goals of elimination as a public health problem and zero transmission, respectively. Robust models, matched to the available data, are the only viable means of quantitatively assessing future dynamics and the long-term impact of controls [8, 17, 18, 21]. Active screening followed by treatment is one of the main control measures, but this action is deployed at the village level suggesting that village-scale models (which recognise the effects of small population size) may be needed to optimise deployment; these results can then be scaled to an infection focus or national level to measure regional elimination, which is especially important as we approach zero transmission.

We have introduced a dynamic, mechanistic, stochastic gHAT model, which is applied to 559 settlements in the Yasa-Bonga and Mosango health zones within the former Bandundu province of the DRC (Fig 1). Using parameters inferred from a deterministic model fitted to aggregated reported cases, our model reliably captures observed detection patterns at the village-scale (Fig 2). This comparison highlighted some spatial heterogeneity associated with the local environmental conditions (significantly fewer zero-detections than predicted occurred in regions close to large rivers, where the tsetse density is presumably high); however, 91.6% of settlements fell within the model (95%) prediction intervals, giving us confidence in our predictive ability. The inclusion of such local environmental factors, which modify the underlying parameters, is clearly an area for further research into refining this small-scale model and may help to practically focus localised control measures, in particular for planning tsetse control.

Throughout our simulation experiments, we consistently find that gHAT persists better in larger populations. This is as expected and agrees with theoretical work and analysis of other diseases [22, 23, 35, 36, 39, 40]; in small populations, the behaviour of the individual is more important, and hence stochastic effects are magnified. The degree of persistence predicted is, however, surprising; settlements of around 2,000 inhabitants, where yearly incidence is only 13 new infections, frequently persist for 15 years or more (Fig 3). This should be contrasted with frequently studied, highly transmissible diseases, such as measles, where local extinctions are common in population sizes of less than 300,000 [22, 23]. We attribute this pronounced difference to the much longer time scales associated with gHAT, meaning single individuals can maintain infection, and the vector-borne nature of gHAT transmission, such that the tsetse act as a short-lived reservoir. We consistently find that incorporating active screening reduces the persistence of infection (Fig 3A), although the distribution of screening across years has only a small effect (Fig 3B). Increasing the screening coverage beyond the average reported levels (of 21% per year) is predicted to lead to further reductions in persistence, but infection is still predicted to be maintained for over 15 years in many larger populations.

Throughout, we have generally ignored the impact of new infectious individuals entering the population, and indeed have shown that this rate of importation is very low. If re-invasion following local elimination is due to the movement of a single infected individual into the settlement (Fig 4B), we predict that the probability of subsequent cases is high (70–80%) and largely independent of population size. However, only a small proportion (10–20%) of such invasions lead to long-term persistence of over 15 years. Re-establishment of infection due to the movement of a limited number of infected tsetse is even less likely. Current uncertainties about the impact of potential reservoirs — either a human reservoir of asymptomatic infections or an animal reservoir — mean there is insufficient knowledge for resurgence to be explicitly modelled by these mechanisms [42], but an animal reservoir that can maintain infection in the absence of humans is likely to represent a worst-case scenario.

A key question, as we approach the 2030 goal of zero transmission, is to ascertain when local elimination has been achieved, allowing policy-makers to scale back control if the infection is no longer present. Due to only a limited proportion of each settlement being screened and the potential for false negatives, a screening can fail to detect any cases even when there is infection in the population. We have shown that, while a single zero-detection screening provides relatively little information of the probability of local gHAT elimination, multiple consecutive zero-detection screenings are a strong indicator of elimination (Fig 5). This can be further strengthened if only large screens (> 50%) are included in the analysis (S1 Appendix, Fig S12B), providing valuable public health information. This concurs with WHO guidelines for active screenings, as villages are no longer considered in planning by mobile screening teams after three consecutive years of zero-detections, followed by a further zero-detection after three years [32]; our model would predict local elimination with large probability for this level of surveillance.

Consistent with the observed patterns in high endemicity regions, we have assumed that the screening coverage and frequency remains constant over these time scales but note that in other regions it may be important to consider the reduction in active screening as reported gHAT cases decline and elimination is approached.

## Conclusion

The ability to capture the stochastic dynamics and persistence of *gambiense* gHAT infection at the village-scale is a major advance in public health modelling, with far-reaching consequences for informing policy decisions. This is particularly pertinent as our models operate at the same spatial scale as controls and can capture the local elimination of infection that is a prerequisite of achieving the 2030 goal of zero transmission globally.

## Supporting information

**S1 Appendix. Model details and supplementary results**. Additional information on the formulation of the model, details of the data and supplementary results.

### Model

#### Model equations

We introduce a compartmental infection model for HAT. The outputs of the model are the number of humans and the proportion of the total number of vectors in each class. For humans, the classes are: susceptible, exposed (but not infectious), Stage 1 infection, Stage 2 infection and hospitalised (or recovering at home). These variables are denoted by *S*_*Hj*_(*t*), *E*_*Hj*_(*t*), *I*_1*Hj*_(*t*), *I*_2*Hj*_(*t*) and *R*_*Hj*_(*t*), where *j* = 1, 2, with *j* = 1 for low risk individuals, randomly participating in active screening, and *j* = 2 for high risk individuals, never participating in active screening, each at time *t* > 0. The population size is assumed to be constant and thus

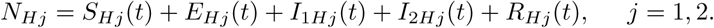

**Fig S1.**
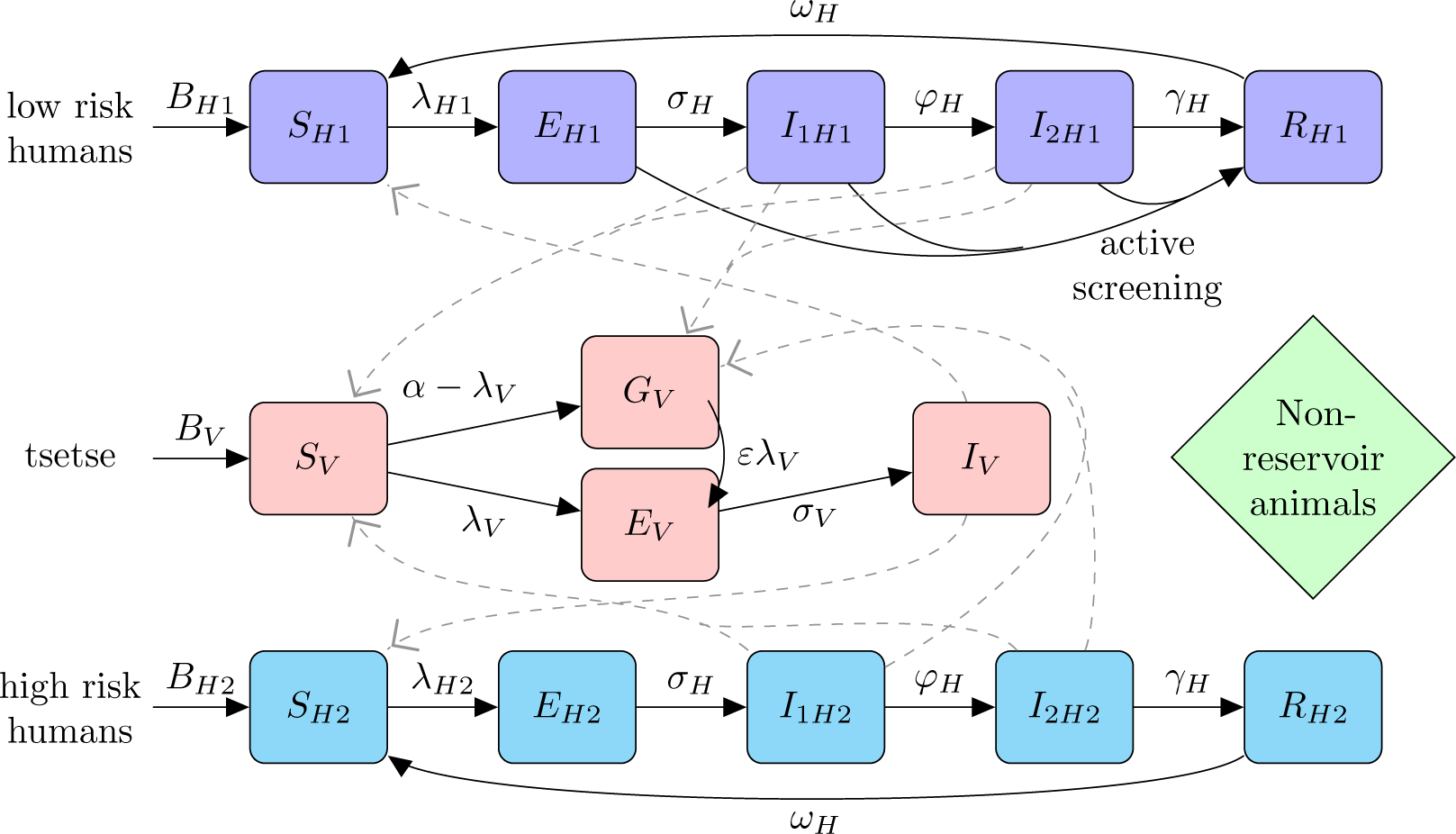
Compartmental model of HAT infection dynamics in humans (low and high risk) and tsetse. Humans can become exposed, *E*_*H*_, on a bite from an infective tsetse and progress through the infection stages, *I*_*H*1_ and *I*_*H*2_, before moving to the non-infectious class *R*_*H*_ due to hospitalisation. Active screening moves exposed or infected people directly to the hospitalised class. Unfed tsetse can become exposed and infected, *E*_*V*_ and *I*_*V*_, on consumption of a blood-meal, of which a proportion are taken on humans. Alternatively, first blood-meals not resulting in exposure means tsetse are less susceptible to trypanosomes in future meals, *G*_*V*_, where we define 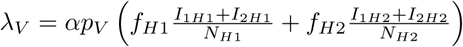. The transmission of infection between humans and tsetse is shown by grey paths. Infected animals are not considered.

The vectors (tsetse) are given by the variables *S*_*V*_, *E*_1*V*_, *E*_2*V*_, *E*_3*V*_, *I*_*V*_ and *G*_*V*_, which correspond to teneral (unfed) tsetse, infected tsetse in their extrinsic incubation period (there are three classes to create a gamma distributed period), infectious tsetse, and non-teneral (fed) but uninfected tsetse. The compartments of the model and possible transitions between them are shown graphically in Fig. S1. We assume that there is natural mortality from all compartments, which leads to replacement of that individual as a susceptible in the population.

Active screening has the potential to detect infection and so move individuals from the exposed or infected classes directly to hospitalised. This is simulated by randomly selecting from the low risk population and if these individuals are exposed or infected, they become hospitalised with 91% probability (the sensitivity of the screening algorithm), as this is the probability of a positive test result, given the person has the infection.

For *m* = *m*_eff_*/p*_*H*_ and *p*_*H*_ > 0, Table S1 defines the transition rates of the Markov-chain model *{*(*S*_*H*1_(*t*), *E*_*H*1_(*t*), *I*_1*H*1_(*t*), *I*_2*H*1_(*t*), *R*_*H*1_(*t*), *S*_*H*2_(*t*), *E*_*H*2_(*t*), *I*_1*H*2_(*t*), *I*_2*H*2_(*t*), *R*_*H*2_(*t*), *S*_*V*_ (*t*), *E*_1*V*_ (*t*), *E*_2*V*_ (*t*), *E*_3*V*_ (*t*), *I*_*V*_ (*t*), *G*_*V*_ (*t*)) : *t* > 0*}* from state (*s*_*H*1_, *e*_*H*1_, *i*_1*H*1_, *i*_2*H*1_, *r*_*H*1_, *s*_*H*2_, *e*_*H*2_, *i*_1*H*2_, *i*_2*H*2_, *r*_*H*2_, *s*_*V*_, *e*_1*V*_, *e*_2*V*_, *e*_3*V*_, *i*_*V*_, *g*_*V*_).

For *m* → *∞* and *p*_*H*_ → 0 (when the vector-to-host ratio, V:H → *∞*), the transition rates remain the same in the human population, but now, since we are considering an infinite number of vectors, an ordinary differential equation approximation is used to describe the vectors. In simulating the model, the tau-leaping algorithm is used, whereby the states and rates are updated with a fixed time step at which the ordinary differential equations are also updated.

In our analysis, the endemic disease equilibrium is calculated as the steady state of the deterministic model, excluding control measures other than basic passive surveillance [1]. Initial conditions for individual villages are taken by sampling from a binomial distribution, where the probability of being selected in a class is given by the proportion in that class in the steady state of the model. Using random initial conditions with the mean at endemic equilibrium removes the effects of rounding the initial conditions to the same integer values for each simulation.

A sample realisation of the infection dynamics in a population with three active screening events, where randomly sampled infected low risk individuals are moved directly into the hospitalised class, is shown in Figure S2.

#### Parameters

Table S2 defines the parameters used in the model for HAT infection dynamics. These parameter values are taken from Rock et al. [1], which were either sourced from literature, where well-defined, or otherwise (in the case of *m*_eff_, *R*_1_, *R*_2_, *r* and *u*) taken as the median of the distribution obtained by model fitting using a Metropolis–Hastings MCMC algorithm that matched the deterministic version of the model to incidence data from the WHO HAT Atlas.

Since the human population is partitioned into low and high risk individuals, we also denote *N*_*H*1_ = *N*_*H*_ *R*_1_ and *N*_*H*2_ = *N*_*H*_ *R*_2_, where *R*_1_ + *R*_2_ = 1. Similarly, *f*_*H*1_ = *f*_*H*_ *R*_1_*/*(*R*_1_ + *rR*_2_) and *f*_*H*2_ = *f*_*H*_ *rR*_2_*/*(*R*_1_ + *rR*_2_) to give the proportion of blood meals on each of the two risk groups, which sum to the total proportion of blood meals tsetse take on humans, *f*_*H*_. Furthermore, we note that the effective density of tsetse is given by the product of the relative tsetse density and the probability of human infection per single infective bite, *m*_eff_ = *mp*_*H*_.

#### Risk structure

The risk structure in the human population is included, since there is evidence for differences in screening attendance and tsetse exposure within the population [3]. Furthermore, using deviance information criterion (DIC) as the method of model selection, there was strong evidence for the inclusion of risk structure [1]. Figure S3 shows the comparison of the models with the reported incidence data. The marginal improvement of the fit when an animal reservoir was added to the model meant animals were not considered here. The DIC was calculated by the following:

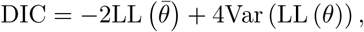

where LL is the log-likelihood of unknown parameters *θ*, and where 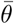 is the expectation of these parameters.

**Table S1.** Model formulation. The transition rates of the Markov-chain gHAT-infection model and additional ordinary differential equation component of the model when *m* → *∞* and *p*_*H*_ → 0 (V:H → *∞*).

| Class | Event | Transition | Rate |
| --- | --- | --- | --- |
| Humans, $j = 1, 2$ | Recovery from hospitalisation | $s_{Hj} \rightarrow s_{Hj} + 1, r_{Hj} \rightarrow r_{Hj} - 1$ | $\omega_H r_{Hj}$ |
| | Natural death of hospitalised | $s_{Hj} \rightarrow s_{Hj} + 1, r_{Hj} \rightarrow r_{Hj} - 1$ | $r_{Hj}$ |
| | Exposure of susceptibles | $s_{Hj} \rightarrow s_{Hj} - 1, e_{Hj} \rightarrow e_{Hj} + 1$ | $f_{Hj} \alpha p_H s_{Hj} i_V / N_{Hj}, p_H > 0$<br>$f_{Hj} \alpha m_{\text{eff}} s_{Hj} I_V / N_{Hj}, p_H \rightarrow 0$ |
| | Progression to Stage 1 infection | $e_{Hj} \rightarrow e_{Hj} - 1, i_{1Hj} \rightarrow i_{1Hj} + 1$ | $\sigma_H e_{Hj}$ |
| | Natural death of exposed | $s_{Hj} \rightarrow s_{Hj} + 1, e_{Hj} \rightarrow e_{Hj} - 1$ | $\mu_H e_{Hj}$ |
| | Progression to Stage 2 infection | $i_{1Hj} \rightarrow i_{1Hj} - 1, i_{2Hj} \rightarrow i_{2Hj} + 1$ | $\phi_H i_{1Hj}$ |
| | Natural death of Stage 1 infection | $s_{Hj} \rightarrow s_{Hj} + 1, i_{1Hj} \rightarrow i_{1Hj} - 1$ | $\mu_H i_{1Hj}$ |
| | Treatment or death from Stage 2 infection | $i_{2Hj} \rightarrow i_{2Hj} - 1, r_{Hj} \rightarrow r_{Hj} + 1$ | $\gamma_H i_{2Hj}$ |
| | Natural death of Stage 2 infection | $s_{Hj} \rightarrow s_{Hj} + 1, i_{2Hj} \rightarrow i_{2Hj} - 1$ | $\mu_H i_{2Hj}$ |
| | Infection importation | $s_{Hj} \rightarrow s_{Hj} - 1, e_{Hj} \rightarrow e_{Hj} + 1$ | $\delta s_{Hj}$ |
| Tsetse ($m = m_{\text{eff}} / p_H, p_H > 0$) | Exposure of teneral | $s_V \rightarrow s_V - 1, e_{1V} \rightarrow e_{1V} + 1$ | $\alpha p_V (f_{H1} (i_{1H1} + i_{2H1}) / N_{H1} + f_{H2} (i_{1H2} + i_{2H2}) / N_{H2}) s_V$ |
| | Exposure of non-teneral, uninfected | $e_{1V} \rightarrow e_{1V} + 1, g_V \rightarrow g_V - 1$ | $\alpha p_V (f_{H1} (i_{1H1} + i_{2H1}) / N_{H1} + f_{H2} (i_{1H2} + i_{2H2}) / N_{H2}) \varepsilon g_V$ |
| | Progression to exposed 2 | $e_{1V} \rightarrow e_{1V} - 1, e_{2V} \rightarrow e_{2V} + 1$ | $3\sigma_V e_{1V}$ |
| | Death in exposed 1 | $s_V \rightarrow s_V + 1, e_{1V} \rightarrow e_{1V} - 1$ | $\mu_V e_{1V}$ |
| | Progression to exposed 3 | $e_{2V} \rightarrow e_{2V} - 1, e_{3V} \rightarrow e_{3V} + 1$ | $3\sigma_V e_{2V}$ |
| | Death in exposed 2 | $s_V \rightarrow s_V + 1, e_{2V} \rightarrow e_{2V} - 1$ | $\mu_V e_{2V}$ |
| | Progression to infected | $e_{3V} \rightarrow e_{3V} - 1, i_V \rightarrow i_V + 1$ | $3\sigma_V e_{3V}$ |
| | Death in exposed 3 | $s_V \rightarrow s_V + 1, e_{3V} \rightarrow e_{3V} - 1$ | $\mu_V e_{3V}$ |
| | Death in infected | $s_V \rightarrow s_V + 1, i_V \rightarrow i_V - 1$ | $\mu_V i_V$ |
| | Fed but not exposed | $s_V \rightarrow s_V - 1, g_V \rightarrow g_V + 1$ | $\alpha (1 - p_V (f_{H1} (i_{1H1} + i_{2H1}) / N_{H1} + f_{H2} (i_{1H2} + i_{2H2}) / N_{H2})) s_V$ |
| | Death in non-teneral uninfected | $s_V \rightarrow s_V + 1, g_V \rightarrow g_V - 1$ | $\mu_V g_V$ |
| Tsetse ($m \rightarrow \infty, p_H \rightarrow 0$) | | $\frac{dS_V}{dt} = \mu_V N_H - \alpha S_V - \mu_V S_V$ | |
| | | $\frac{dE_{1V}}{dt} = \alpha p_V \left( f_{H1} \frac{I_{1H1} + I_{2H1}}{N_{H1}} + f_{H2} \frac{I_{1H2} + I_{2H2}}{N_{H2}} \right) (S_V + \varepsilon G_V) - (3\sigma_V + \mu_V) E_{1V}$ | |
| | | $\frac{dE_{2V}}{dt} = 3\sigma_V E_{1V} - (3\sigma_V + \mu_V) E_{2V}$ | |
| | | $\frac{dE_{3V}}{dt} = 3\sigma_V E_{2V} - (3\sigma_V + \mu_V) E_{3V}$ | |
| | | $\frac{dI_V}{dt} = 3\sigma_V E_{3V} - \mu_V I_V$ | |
| | | $\frac{dG_V}{dt} = \alpha \left( 1 - p_V \left( f_{H1} \frac{I_{1H1} + I_{2H1}}{N_{H1}} + f_{H2} \frac{I_{1H2} + I_{2H2}}{N_{H2}} \right) \right) S_V - \alpha p_V \left( f_{H1} \frac{I_{1H1} + I_{2H1}}{N_{H1}} + f_{H2} \frac{I_{1H2} + I_{2H2}}{N_{H2}} \right) \varepsilon G_V - \mu_V G_V$ | |

**Fig S2.**
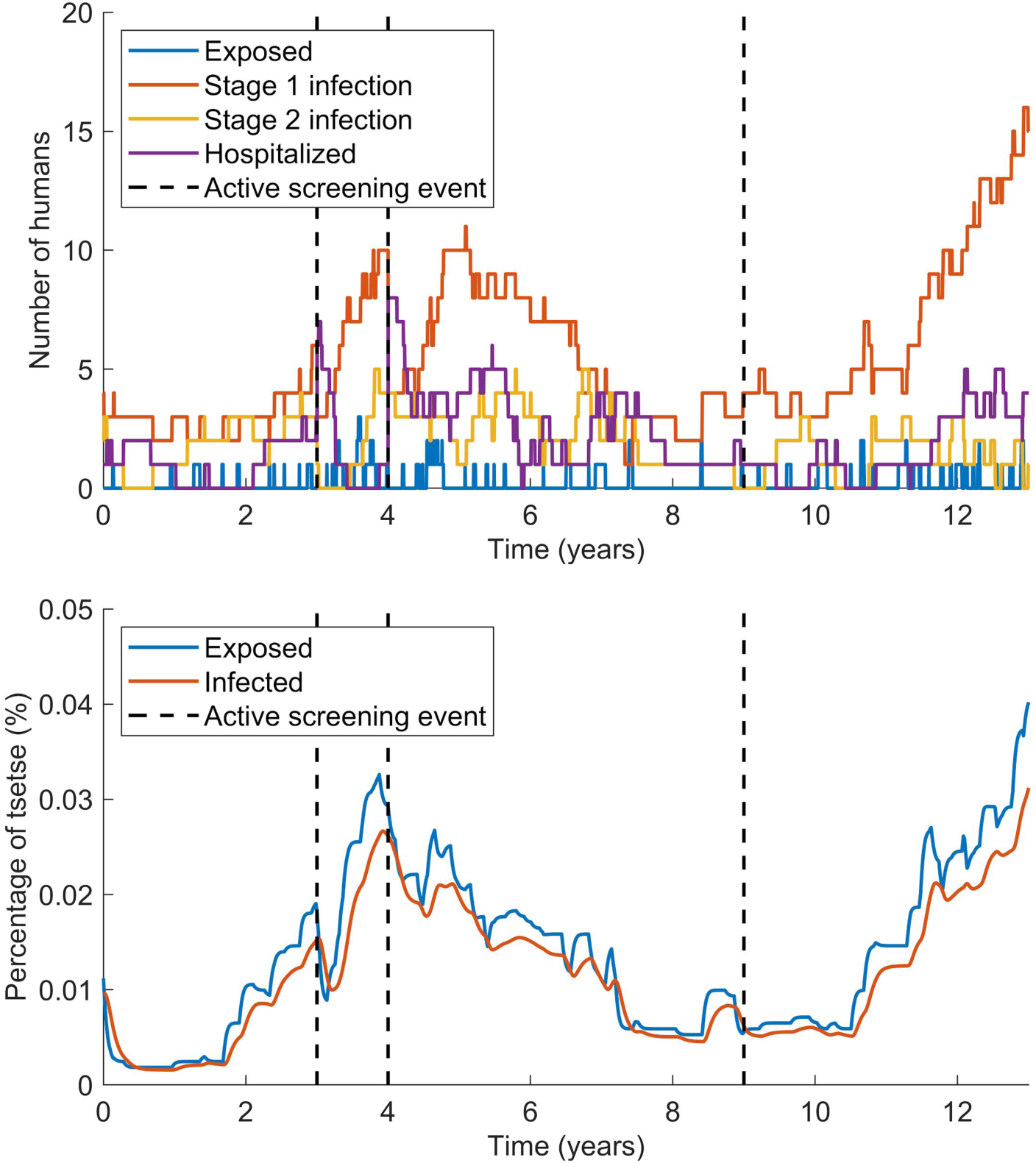
A single realisation of the simulated infection dynamics of the model. A population of 514 is used with 404, 430 and 52 of the low risk population randomly selected for active screening after three, four and nine years, respectively. Note that the number of hospitalised individuals increases at the first two active screening events, but no infected individuals are randomly selected and detected in the third screening event. We only plot those infected (or affected) by HAT, although we note that the vast majority of both populations are susceptible.

**Table S2.** Model parameters. Parameter notation and values for the stochastic model.

| Parameter | Description | Value | Source |
| --- | --- | --- | --- |
| $\mu_H$ | Natural human mortality rate | $5.4795 \times 10^{-5} \text{ days}^{-1}$ | [2] |
| $\omega_H$ | Human recovery rate | $0.006 \text{ days}^{-1}$ | [3] |
| $\sigma_H$ | Human incubation rate | $0.0833 \text{ days}^{-1}$ | [4] |
| $\phi_H$ | Stage 1 to 2 progression rate | $0.0019 \text{ days}^{-1}$ | [5,6] |
| $\gamma_H$ | Exit rate from Stage 2 by treatment or death | $0.006 \text{ days}^{-1}$ | [1] |
| $N_H$ | Human population size | Varies | N/A |
| $\mu_V$ | Tsetse mortality rate | $0.03 \text{ days}^{-1}$ | [4] |
| $\sigma_V$ | Tsetse incubation rate | $0.034 \text{ days}^{-1}$ | [7,8] |
| $\varepsilon$ | Reduced non-teneral susceptibility factor | 0.05 | [1] |
| $\alpha$ | Tsetse bite rate | $0.333 \text{ days}^{-1}$ | [9] |
| $m$ | Relative tsetse density | Varies | N/A |
| $p_H$ | Probability of human infection per single infective bite | Varies | N/A |
| $m_{\text{eff}}$ | Effective tsetse density | 6.56 | [1] |
| $p_V$ | Probability of tsetse infection per single infective bite | 0.065 | [4] |
| $f_H$ | Proportion of blood-meals on humans | 0.09 | [10] |
| $R_1$ | Proportion of low risk humans, randomly participating in screening | 0.924 | [1] |
| $R_2$ | Proportion of high risk humans, not participating in screening | 0.076 | [1] |
| $r$ | Relative bites taken on high risk humans compared to low risk | 6.6 | [1] |
| $u$ | Reporting probability for Stage 2 cases | 0.26 | [1] |
| $\delta$ | Importation of infection rate | $3.4 \times 10^{-6} \text{ days}^{-1}$ | fitted |

**Fig S3.**
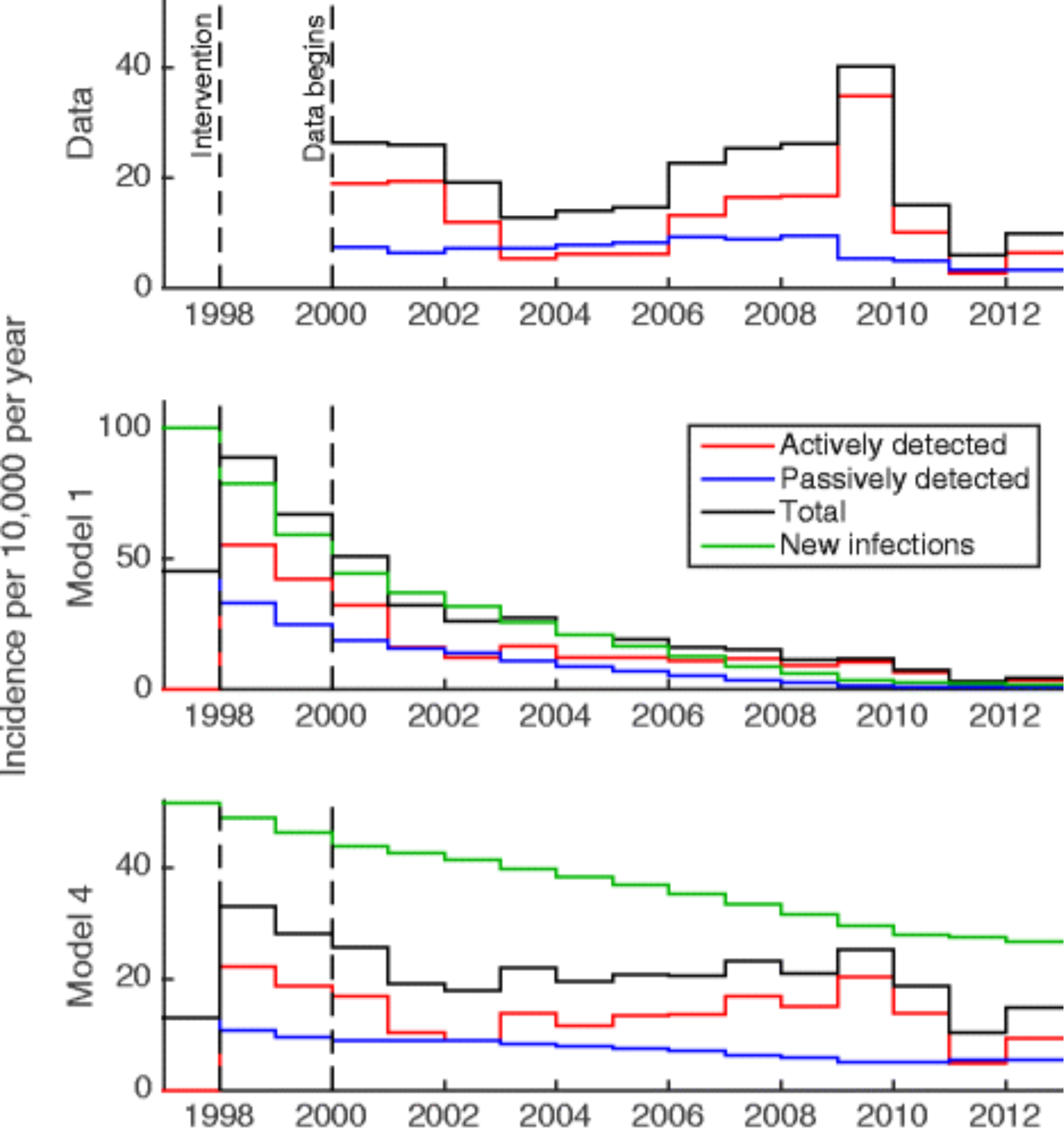
Deterministic model selection. Comparison of the reported incidence data aggregated across the study region and the corresponding model output over the years 1998–2012, for the deterministic model, both without risk structure (Model 1) and with risk structure (Model 4). Model 4 provides the better fit with the incidence data. This figure is adapted from Rock et al. [1].

#### Tsetse density

One fundamental parameter of the model, the effective density of tsetse, is equal to the product of the vector-to-host ratio and the probability of human infection per single infective bite. In the original deterministic framework, these constituent terms did not need to be known, as it was only the product that was important; therefore they could not be inferred separately. However, since the stochastic model explicitly captures the number of tsetse, the two parameters are now required. Given the limited data to estimate either component parameter, we choose to explore the potential range of parameters: from very high probability of infection and hence low vector-to-host ratio, to very low probability of infection and large vector-to-host ratio such that the dynamics of tsetse can be modelled deterministically.

Considering the simplest case in the absence of disease-control measures, we find that different values for the vector-to-host ratio have a relatively limited impact on predictions of disease persistence. Unsurprisingly, large tsetse populations, which can be approximated by deterministic dynamics, lead to the greatest persistence (Fig. S4); thus, throughout this paper, we utilise this worst-case scenario, although other assumptions do not change the qualitative findings.

**Fig S4.**
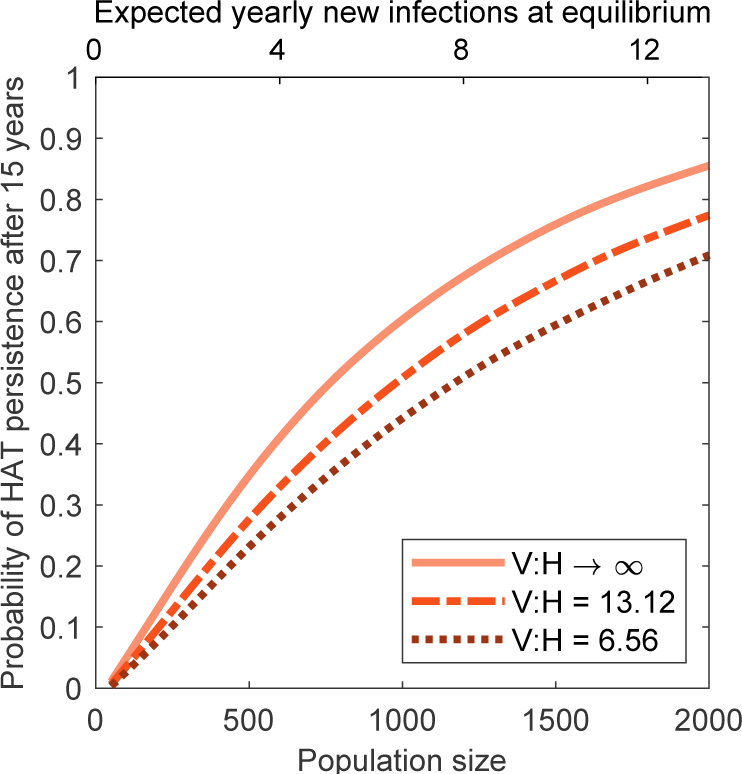
Impact of the parameters forming the effective density of tsetse. When the probability of human infection per single infective bite is high the vector-to-host ratio, V:H, is around 6.56, whereas when the probability is low, V:H becomes large and deterministic models are valid for tsetse dynamics. The expected number of yearly new infections for each population size, if the system were at equilibrium, is also given by the top scale.

#### Isolated populations

For the majority of results in the main paper, we assume that importations of infection between villages do not play a significant role and so treat them independently. In many cases, such as community-level persistence, this approach is justified, as we wish to understand the stochastic dynamics in the absence of confounding imports. We also quantify the level of infectious imports by considering the probability of a village having infection present on the first screening and assuming this represents a long-term equilibrium solution, since this is the infection level before regular active screening began. By matching the WHO HAT Atlas data to the output of the model with an additional per person importation rate parameter (*δ*) we find that, using least-squares fitting, *δ*≈ 3.4 × 10^−6^ days^−1^. This rate is small and so can be reasonably ignored on the studied time scales, particularly as there is some probability that this importation will not cause any subsequent transmission. In addition, it is reasonable to assume that this importation rate will decay over time, since the total case numbers in the DRC is reducing [11]. By fitting the reduction in total cases in the DRC to an exponential decay function, we get a decay rate of 0.1071, which we apply to the import rate (Fig. S5).

**Fig S5.**
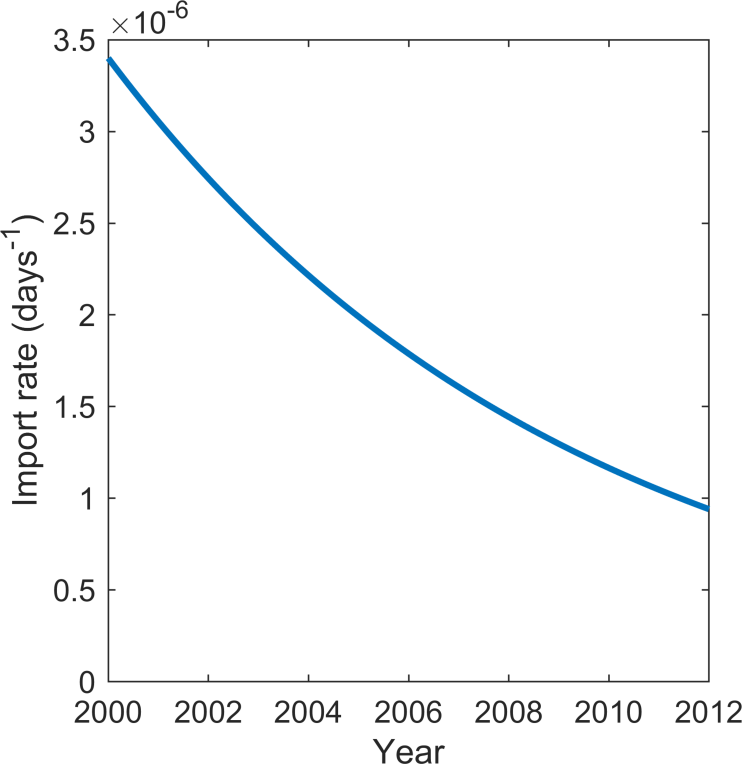
Change in the importation of infection parameter in time. The decay of the rate of importation of infection parameter is matched to the decay rate of cases of HAT in the DRC [11].

As a validation of our assumption that importations can be safely ignored over the time-scales of this study, we re-examine the match between active-screenings that fail to detect cases and model results including imports (Fig. S6). This is a counter-point to Fig. 2 in the main paper. We find that the inclusion of imports has an extremely small difference on the model predictions: slightly improving the fit between model and data (Fig. S6A). However, under this new model, there is now an additional village where the model significantly predicts fewer zero-detections than observed, decreasing the percentage of villages within the model prediction intervals to 91.4% (Fig. S6B).

**Fig S6.**
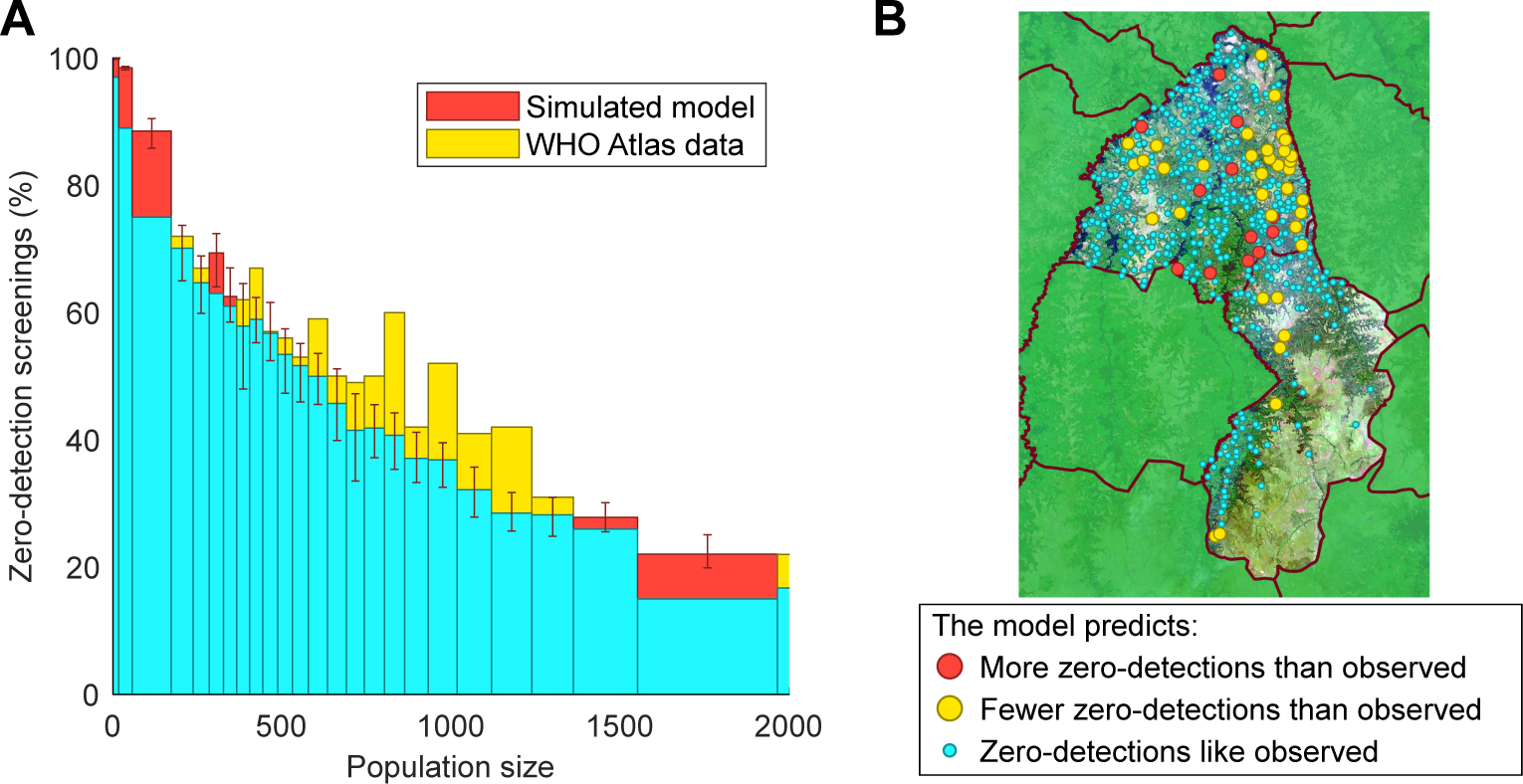
Comparison of model predictions and data for active screenings with no detected cases (zero-detections) for the model with importations of infection. (A) Histogram by population size of the percentage of active screenings that find no new HAT cases for both the model and the HAT Atlas data. The differences between this histogram and without imporatations of infection are very small. (B) Map of populations in Yasa-Bonga and Mosango showing the settlements with significant differences (at the 95% level) in the expected proportion of active screenings with no cases detected. The addition of importations of infection means this model fits the data less well, with an additional village falling outside the prediction intervals. The underlying Landsat-8 satellite map is shown courtesy of the U.S. Geological Survey.

#### Tau-leaping

Since we use a fixed step size for the tau-leaping algorithm of 0.1 days, Figure S7 shows that the results for persistence of HAT after 15 years are qualitatively the same for time steps of 0.01 and 1 days, so we are confident that a time step of 0.1 days is not introducing numerical errors. 95% confidence intervals are all small, in the range (0.0044, 0.0196) about the plotted mean values.

**Fig S7.**
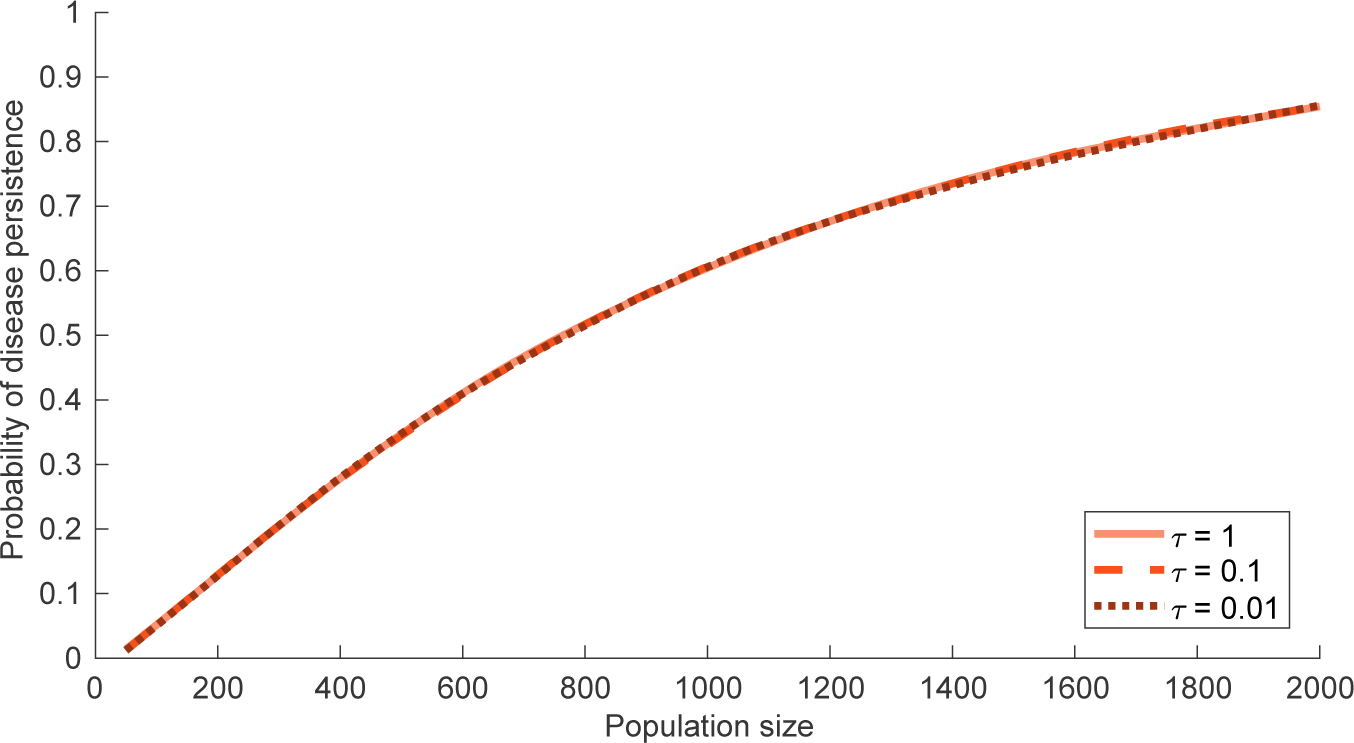
Probability of disease persistence under the tau-leaping algorithm for fixed step size. Probability of HAT persistence after 15 years for V:H→ *∞*, simulated using three different tau-leaping time steps: *τ* = 0.01, 0.1 and 1. Since the resulting curves are the same, the step size in the algorithm is suitable.

### Data

#### Active screening data

In the main manuscript, we note that not all settlements undergo the same levels of screening. In fact, larger village populations tend to have more frequent screening, but with a lower proportion of the population tested; small populations tend to have high coverage but infrequent screens (Fig. S8A and B). The smaller villages were also not typically screened in 2000–2008, while villages with populations greater than 500 were screened over the whole data period 2000–2012 (Fig. S8C). Typically, we see that 50% of villages were screened in any given year. We do not account for any of these differences in our analysis but can use these trends to help explain any discrepancies between models and the data.

In addition, the active screening data from the WHO HAT Atlas were used to calculate the mean annual screening coverage as 21%. Population sizes were taken from the census data within the WHO HAT Atlas and then modified by assuming an annual population growth of 2.6%. Coverages were then calculated as the number of people screened divided by the estimated population size, assuming a maximum coverage equal to that of the entire the low risk population size (92.4%), as described in the main manuscript. The mean annual coverage was then obtained as the mean of these coverages, where active screening occurred, as well as the 0% coverages, where no active screening was recorded. This value is thus highly dependent on the quality of the census information, as differences in population sizes will change this estimate of mean coverage. It is therefore important to have good census data for making predictions on the effect of different screening coverages.

**Fig S8.**
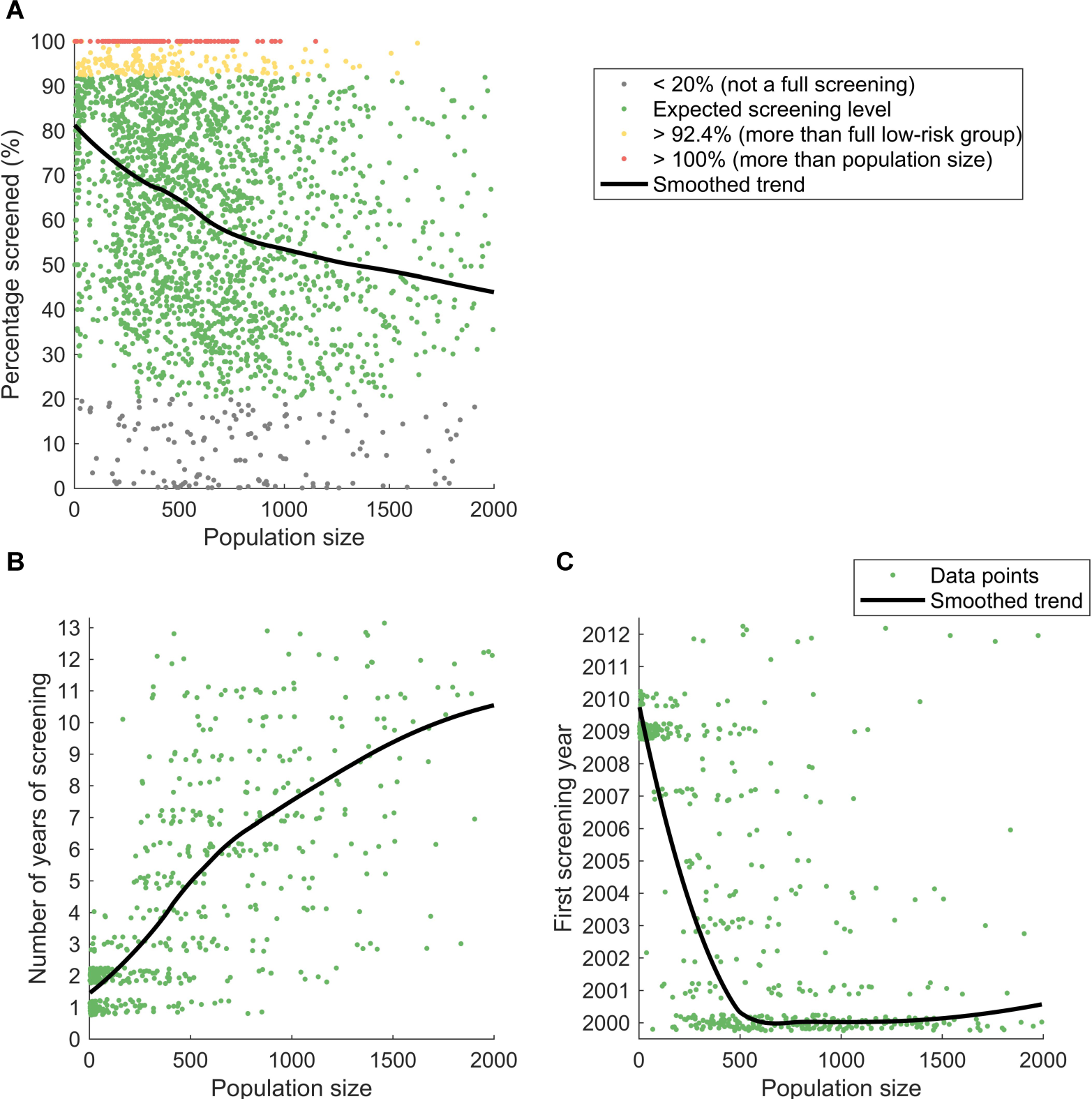
Differences in screening between villages. (A) Screening coverage in village populations. (B) Number of screenings at each settlement in the period 2000–2012. Small random perturbations are made to the integer values for visibility. (C) Years in which the first active screenings occur in the villages (within the 2000–2012 period), with small random perturbations for visibility.

### Persistence and elimination

#### Local disease persistence

As a control strategy, we find that randomly selecting the screening coverage from all reported coverages slightly out-performs screening annually at the mean coverage (21%), kept fixed across all years. High levels of coverage in any one year are more likely to interrupt transmission (Fig. S9A). In addition, long-term persistence requires continued transmission and so the probability of persistence decreases with time (Fig. S9B and C).

**Fig S9.**
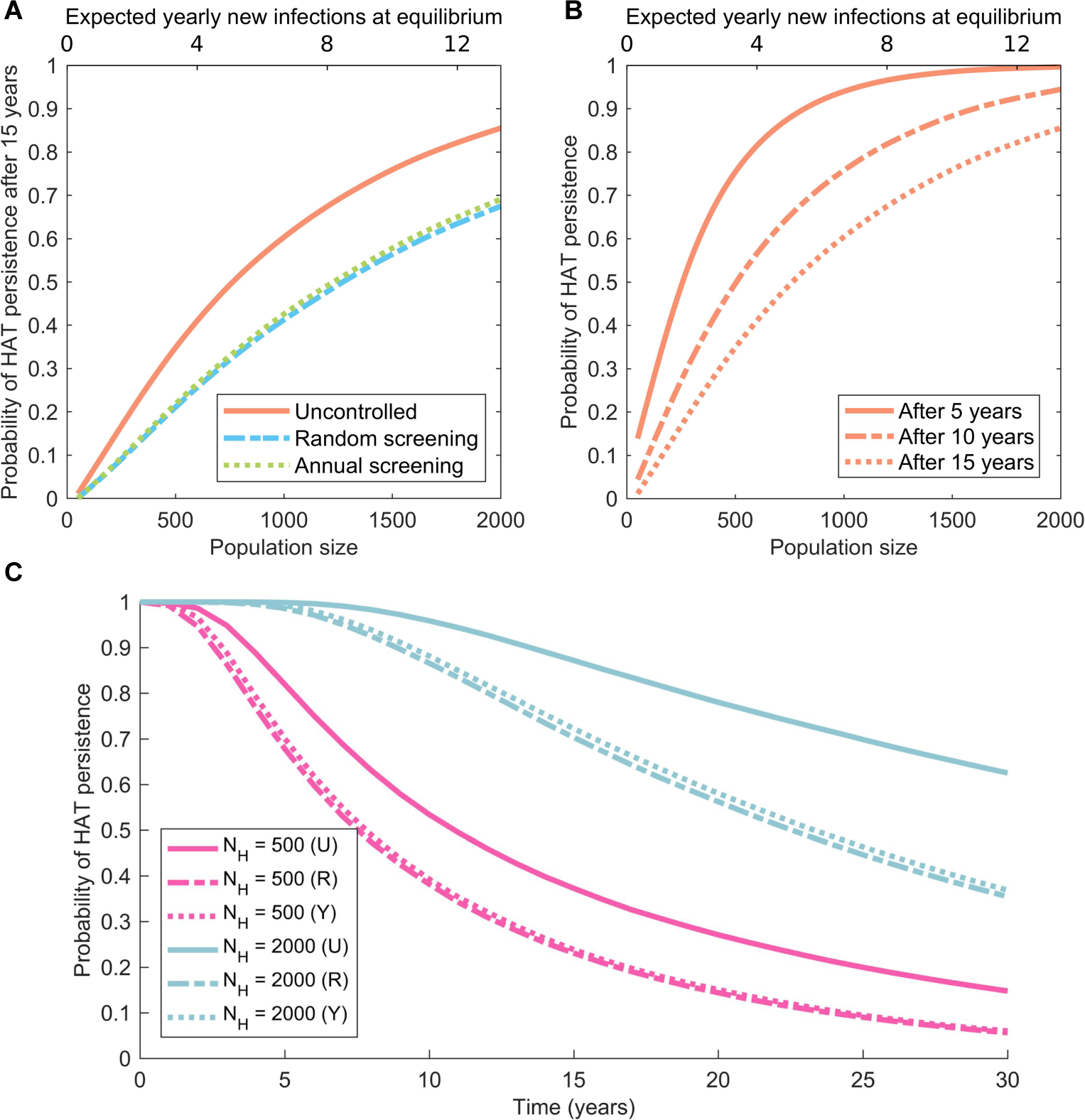
Factors affecting predicted probability of HAT persistence in isolated settlements. (A) Impact of active screening on disease persistence: both random screening (sampling from all screening coverages, including zeros) and annual screening at a fixed coverage (the mean annual coverage of 21%) yield a drop in persistence. Choosing the coverage randomly slightly out-performs screening annually at the mean coverage. (B) Probability of HAT persistence 5, 10 and 15 years after starting at the endemic equilibrium. (C) For villages with populations of sizes 500 and 2,000, the probability of HAT persistence in time for the uncontrolled setting (U), random active screening coverage (R) and annual screening at the average coverage of 21% (Y).

#### Re-invasion due to tsetse

We consider the dynamics of re-invasion, given different initial conditions to Figure 4 in the main manuscript. In a population without human infections but with endemic levels of tsetse infection, the chance that any human cases will be generated increases with settlement size (Fig. S10A). This is due to tsetse being able to interact with fewer humans over their short lifespan, while new transmission events caused by human infection are less dependent on population size, as the infection will be present in the population for much longer. The probability of new transmission events is further increased by the addition of an infected human to endemic levels of tsetse infection (Fig. S10B).

**Fig S10.**
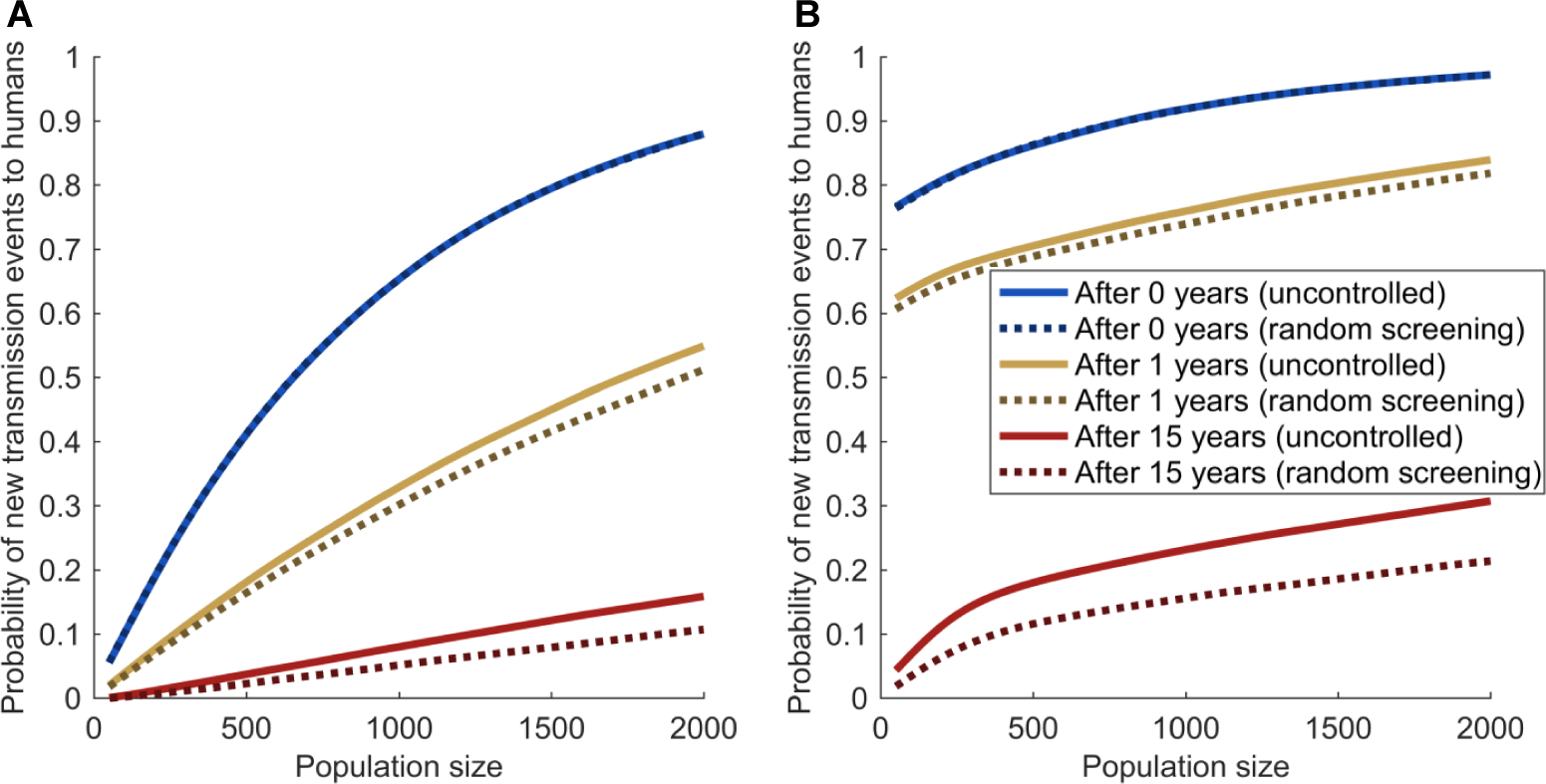
Dynamics of re-invasion. The probability that at least one human case is generated, and cases continue to be generated beyond for a given times. (A) Starting with no human infection and endemic-equilibrium levels of infected tsetse. (B) Starting with one infected human and endemic-equilibrium levels of infected tsetse.

#### Proportion of positive tests in a screening

Comparing the data on the proportion of tests that result in a positive detection of HAT to the output of the model demonstrates similar results to comparing the screenings with no detected cases, as in Figure 2 from the main manuscript (Fig. S11). The difference in coverages of an active screening event mean the results have more variation than simply not detecting any new cases, yet the model is similarly capturing the behaviour seen in the data; there are a few more settlements that stand out as significantly different from the model (69 settlements, 12.3%), but these are similarly clustered where expected.

#### Definition of a full screen

In calculating the probability of infection on detecting no cases, a zero-detection event is different for different screening percentages. We use 20% of the population screened to define a ‘full screening’; however, similar results are achieved for the screening regime of Yasa-Bonga and Mosango 2000–2012 when 10% is used. If 50% is used, and so all screenings are of reasonable quality, there is more confidence that the same number of consecutive zero screenings are a proxy for no infection in the population (Fig. S12).

**Fig S11.**
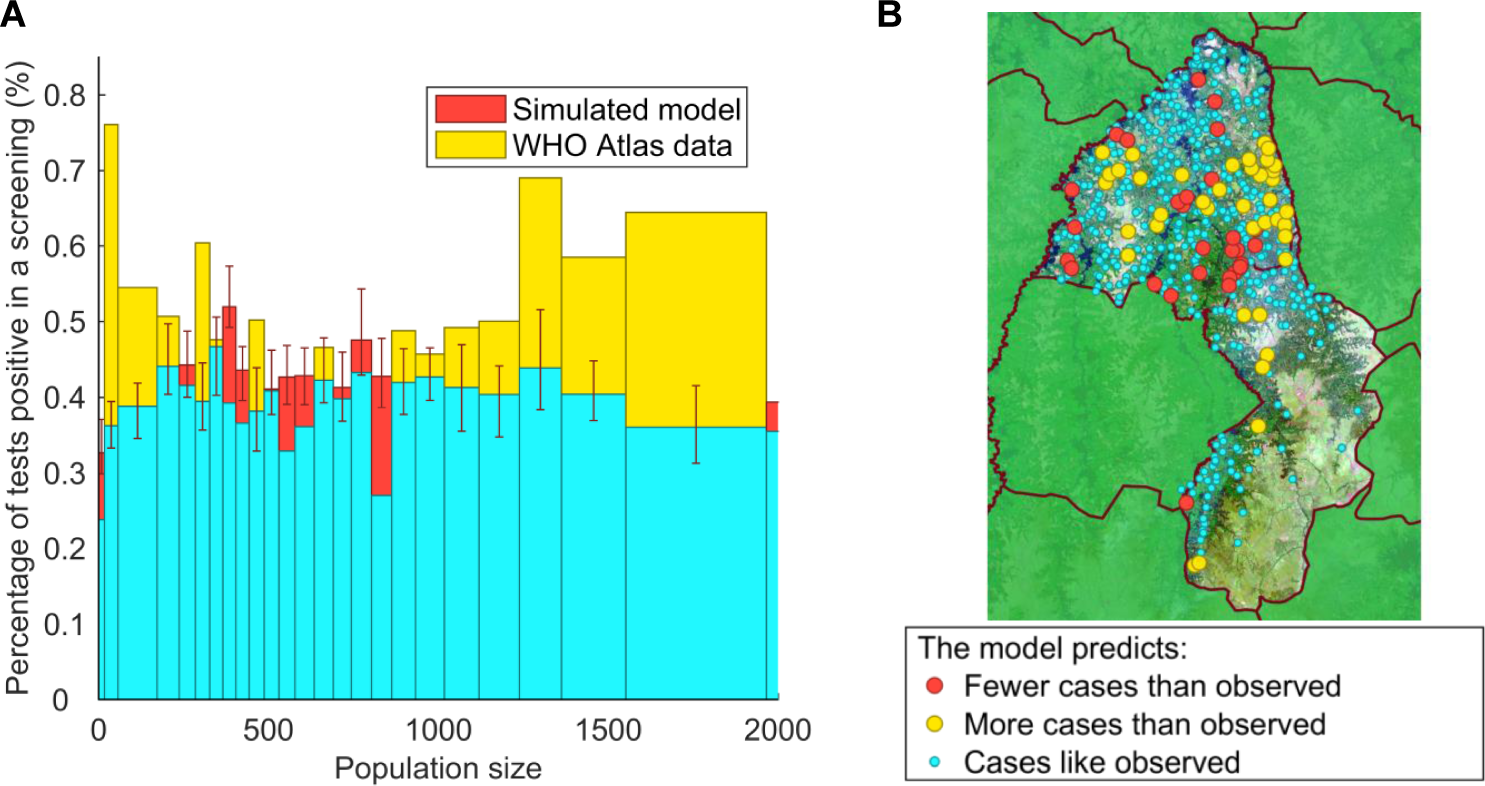
Expected proportion tests that are HAT positive in an active screening. (A) Histogram by population size of the percentage of positive tests in independent active screenings for both the model and the WHO HAT Atlas data. (B) Map of populations in Yasa-Bonga and Mosango showing the settlements with significant differences in the expected proportion of tests positive. The underlying Landsat-8 satellite map is shown courtesy of the U.S. Geological Survey.

When screening coverage is higher and no infection is detected, there is more certainty that no infection is present.

**Fig S12.**
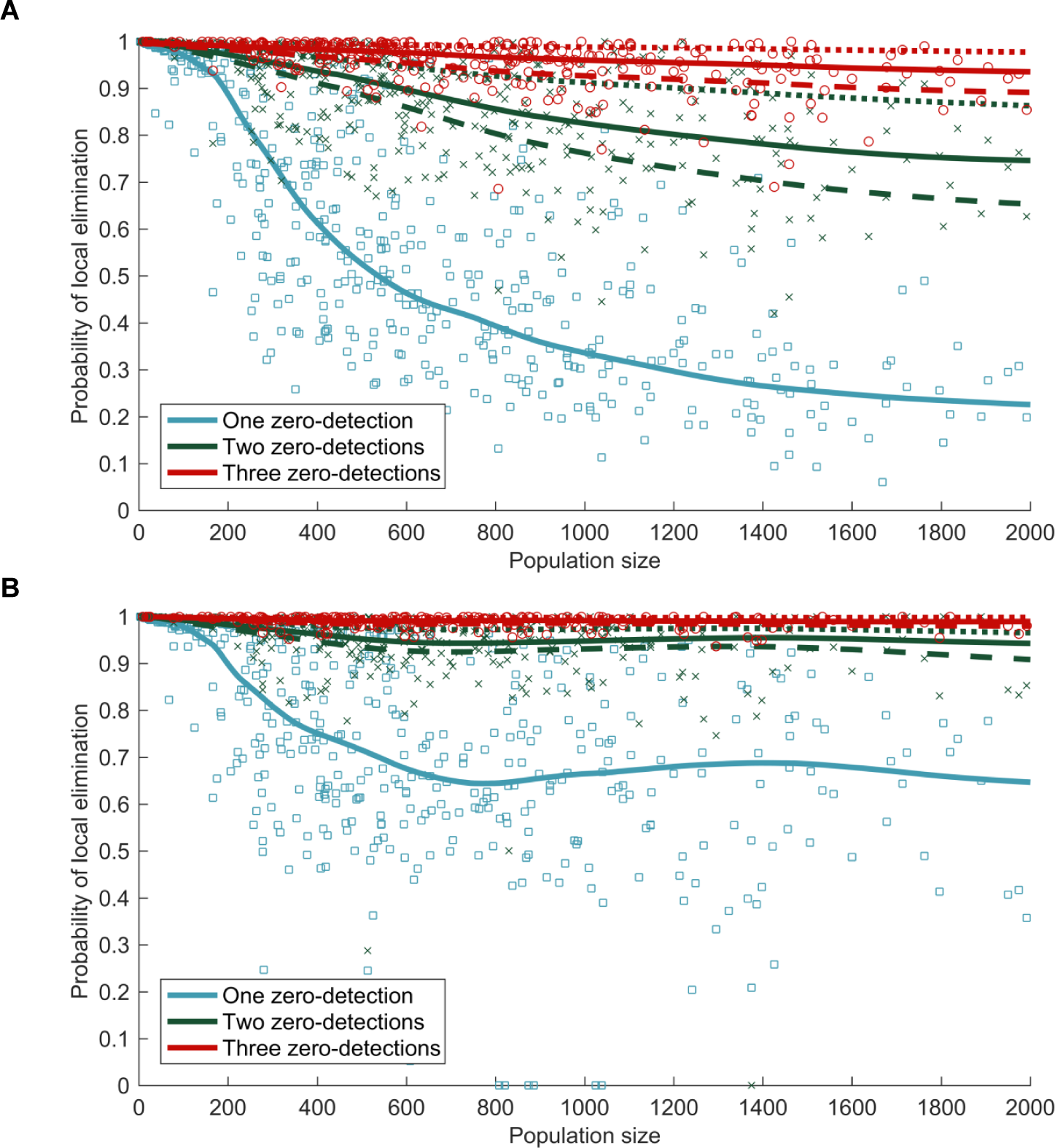
Probability of elimination in a settlement given no cases have been detected in consecutive screenings. The probability of a settlement containing no infection given that a number of consecutive active screenings detecting no cases have occurred. (A) Only active screenings where at least 10% of the population are included are used to calculate the probability. (B) At least 50% of the population must be screened for the screening to be included.

## Data Availability

Epidemiological data for the study were provided by the WHO in the frame of the Atlas of gHAT which may be viewed at www.who.int/trypanosomiasis_african/country/risk_AFRO/en and may be requested through Jose Ramon Franco.

**S1 Model code. C+ + code.** The code to run the basic stochastic model.

## Acknowledgments

This work was supported by the Bill and Melinda Gates Foundation (www.gatesfoundation.org) in partnership with the Task Force for Global Health through the NTD Modelling Consortium [OPP1053230] (K.S.R. and M.J.K.), the Bill and Melinda Gates Foundation through the Human African Trypanosomiasis Modelling and Economic Predictions for Policy (HAT MEPP) project [OPP1177824] (K.S.R. and M.J.K.) and EPSRC/MRC via the MathSys Centre for Doctoral Training (C.N.D.). The views, opinions, assumptions or any other information set out in this article are solely those of the authors and should not be attributed to the funders or any person connected with the funders.

